# The different gene-environment interaction pattern in major depressive disorder and generalised anxiety disorder: a comparative study across 36 environments

**DOI:** 10.1101/2025.11.10.25339885

**Authors:** Qianshu Ma, Yulu Wu, Yunqi Huang, Min Xie, Shiwan Tao, Yuanjun Gu, Siyi Liu, Varun Warrier, Qiang Wang

## Abstract

**Background:** Major depressive disorder (MDD) and generalised anxiety disorder (GAD) are common psychiatric disorders. Previous studies showed shared genetic and environmental factors, but no studies have compared their overlap and differences from gene-environment interaction perspective. Current studies neglect the high rate of comorbidity between them and have overly broad case definitions, which may inflate the shared risk. In this study, we strictly defined the inclusion criteria to define three groups: MDD-only, GAD-only, and comorbid group, to investigate similarities and differences in gene–environment interactions across these groups.

**Methods and materials:** We analysed data from 396,443 participants in the UK Biobank (UKB), after applying exclusion criteria. We divided case group into three groups: MDD-only, GAD-only and comorbid group. 36 environmental variables across the three dimensions (socio-demographic, early life experiences and lifestyle) were included in this study and the PHESANT package was used to find significant environmental variables associated with three phenotypes. Logistic regression models were established with significant environmental variables and polygenic risk scores (PRS) of depression and anxiety as independent variables to test for PRS × environment interaction effects. Finally, the Genome-wide Gene-environment Interaction Association Studies (GWEIS) were performed using fastGWA-GE to identify susceptibility loci interacting with the significant environmental variables, and their biological annotations were conducted by FUMA and MAGMA.

**Results:** In this study, 396,443 participants (53.9% female) were included, the MDD-only (n = 58,582), GAD-only (n = 12,063), comorbid (n = 26,119) and control groups (n = 299,679) differed significantly across all sociodemographic variables (*P*<0.001). Environmental screening identified 19 variables shared by all three groups, along with 2 environmental variables unique to MDD-only, 1 environmental variable unique to the GAD-only. In the GWEIS analyses, SNPs interacted with 5 stressful life events passed the Bonferroni correction to affect MDD-only; 8 SNP-environment interactions passed the Bonferroni correction to significantly affect GAD-only, including childhood maltreatment and recent stress; and 3 environmental variables, including recent stress and adoption, significantly interacted with the SNPs to influence comorbid phenotype. Same SNP and financial crisis interaction pattern was found in all three groups, but the susceptibility SNPs that significantly interacted with financial crisis were different. MR results showed childhood maltreatment causally increased the risk of MDD-only, GAD-only and their comorbidity.

**Conclusion:** Findings indicate MDD-only and GAD-only have more distinct socio-demographic, environmental factors and gene-environment interaction patterns after excluding comorbid cases. There are also difference in socio-demographic and gene-environment interaction patterns when comparing MDD with and without comorbidity. It may provide potential biological evidence for heterogeneity within diseases and clinical intervention for subtypes.

## Introduction

Major depressive disorder (MDD) and generalised anxiety disorder (GAD) are two common psychiatric diseases. The life-time prevalence of MDD and GAD is 6.8% and 7.6 %, respectively^1^. They have shared genetic and environmental factors. Meta-analyses have illustrated that environmental factors influence the onset of these two diseases in three dimensions: socio-demographic, early experiences and lifestyle^2^. Certain socio-demographic features, such as gender, income and education, are significantly associated with MDD and GAD^3^. Early experience including childhood maltreatment, smoking during pregnancy, multiple birth and low weight at birth have been identified as transdiagnostic risk factors for MDD and GAD^4,5^. Lifestyle factors, such as stressful life events, smoking, obesity, substance use and digital device use have also been associated with an increased onset of MDD and GAD^6,7^. Several studies have shown that individuals with depression and anxiety tend to have a higher percentage of experiencing stressful life events, smoking, poorer physical activity levels, substance use and longer digital device use compared to healthy controls^8-11^.

Twin studies have estimated the heritability of MDD and GAD to be 37% and 32%, respectively^12,13^. Early genetic studies selected candidate genes through hypotheses of disease mechanisms, and MDD and GAD were found to share candidate genes such as the serotonin transporter protein gene (5-HTT) and the dopamine receptor gene (DRD2)^14^. Thorp et al found that 509 genes were shared between depressive and anxiety disorder symptoms^15^. Some of the genes identified by recent GWAS studies for GAD can also be found in the MDD GWAS results^16^. Moreover, twin studies showed that MDD and GAD had completely shared genetic factor and the difference in environments decided whether they had MDD or GAD^17^. The genetic correlation between MDD and anxiety disorders was estimated to be 0.91^18^ using GWAS data. The above studies showed that MDD and GAD shared environmental and genetic factors.

Although MDD and GAD have overlap in clinical symptoms, environmental and genetic factors, they belong to different disease spectrums and have different disease courses, core symptoms, and neurological responses, all of which indicate that they are different psychiatric disorders. Thus, we hypothesised that the gene-environment interaction patterns of the two might be different. To investigate the genetic and environmental effects on MDD and GAD, Kendler found that they shared genetic factors exclusively while shared family factors had no effect on the two through twin studies^17^. However, epidemiological studies have suggested that a proportion of mental illness could be partially attributed to environmental factors including poverty and childhood abuse, which are shared within families^4^. This contradiction between these two results is called the shared environment paradox. The emergence of the gene-environment interaction (G×E) hypothesis can provide a simple explanation for this paradox: if the effect of the shared environment is dependent on genetic variation, the statistical models used in twin studies would attribute the joint effect exclusively to genetic contribution, thus exaggerating the proportion of genetic sharing in twin studies^19^. This hypothesis suggests that genetic factors cause individuals to have a predisposition to be sensitive to dangerous environmental factors, and whether an individual develops a mental disorder depends on the dynamic interaction between the two. The theoretical model explains why some people will develop mental disorders while others will not after a population is exposed to a specific risk.

Early studies focused on using candidate strategy genes to explore the effects of specific genes interacting with environmental factors on depression and anxiety^20^. Recent studies have been focusing on exploring the G × E patterns of MDD and GAD, respectively. Genome-wide Gene-environment Interaction Association Studies (GWEIS) have become a popular methodology for investigating gene-environment interaction. There are limited studies using GWEIS to explore gene-environment interaction in mental health. Dunn et al. were the first group to utilize a large-scale genome-wide gene-environment interaction design and found that SNPs significantly interacting with stressful life events had an impact on depressive symptoms^21^. However, to date in the field of psychiatric disorders, GWEIS studies have focused only on a limited number of environmental variables (e.g. childhood maltreatment and stressful life events) and on studies focusing on the symptomatic level (phenotypes such as depression, anxiety and personality disorders)^22,23^.

However, some of the current study designs had limitations. There has been an increasing need for large genetic studies of MDD and GAD because a larger sample size can increase statistical power. However, recruiting cohorts with an enlarged sample size often comes with simple methods (sloppy case definition and ignorance of comorbidity with other psychiatric conditions). As a result, a proportion of identified loci lacked specificity to the genetic contribution to MDD, which may potentially bias the results of MDD genetic etiology^24,25^. Comorbidity between MDD and anxiety disorders is common, with studies showing that 75% of patients with MDD also have a diagnosis of anxiety disorder, of which GAD has the highest comorbidity with MDD^26^. Approximately 59% of patients with GAD also meet the diagnostic criteria for MDD^27^. In clinical practice, MDD patients with GAD often have a worse prognosis, more severe symptoms, an earlier age of first onset of MDD, a poorer quality of life, higher rates of recurrence of MDD, and a higher risk of suicide than patients without GAD^28^. The neglect of depression-anxiety comorbidity in genetic study designs may lead to confusion in the interpretation of results. For example, a GWAS study for GAD identified genes that partially overlapped with the results of a meta-analysis of GWAS for depression^16,29^. However, 53% of the participants in the study had both diagnoses of MDD and GAD, making it difficult to determine whether the overlapping genes were due to depression-anxiety comorbidity in the sample or whether there were shared genes between the two disorders.

Based on the above hypothesis and the shortages of current study designs, we utilized strict inclusion criteria to define three groups: MDD-only, GAD-only, and the comorbid group, in order to compare the gene-environment interactions patterns across these groups. We included 36 environmental variables across three dimensions (socio-demographic, early life experiences and lifestyle) and the PHESANT package was used to find significant environmental variables associated with the three groups. Logistic regression models were established with significant environmental variables and polygenic risk scores (PRS) of depression and anxiety as independent variables to test for PRS × environment interaction effects. Finally, the Genome-wide Gene-environment Interaction Association Studies (GWEIS) were performed on the three groups using fastGWA-GE to identify susceptibility loci interacting with the significant environmental variables. To further find out the causality between environmental risk factors and outcomes, we conducted MR using … Finally, we compared the susceptible loci among three groups to investigate whether MDD and GAD have different gene-environment interaction patterns.

## Methods and Materials

### Participants

The UK Biobank (UKB) cohort was assembled across 22 assessment centres in the UK between 2006 and 2010, recruiting over 500,000 participants aged from 38 to 72^30^. In this study, we used data at baseline and online follow-up in 2016 (mental health) and 2022 (mental well-being). After exclusion, 396,443 participants were included (Figure S1 in the supplementary). The detailed inclusion and exclusion criteria are described below.

### MDD and GAD case definition

Three definitions were included to define cases: 1) the Composite International Diagnostic Interview Short Form (CIDI-SF); 2) the International Classification of Diseases, Tenth Revision (ICD-10) based on electronic medical records; 3) self-reported diagnoses as part of past and current medical conditions. To maximize the sample size, we included participants who fit into any one of the three definitions of MDD and GAD into case groups. We divided them into three subgroups: MDD-only, GAD-only and the comorbid group. These definitions were also used in previous research^31,32^. The detailed case definitions are described in Table S1.

### Control group definition

The control group included participants who did not fit any of these three case definitions and had scores lower than 10 on both the PHQ-9 and GAD-7 scales (Table S1).

### Exclusion for case and control groups

We excluded any case or control who met the criteria of ICD-10 or self-reported schizophrenia, bipolar disorder I or II. A total of 6,188 participants were excluded according to these criteria. Additionally, we excluded any controls who had a score higher than 10 on PHQ-9 or GAD-7 scales or had no record on the scales (Figure S1).

### Genotype data and quality control

All genotype data were obtained from the UK Biobank. Detailed primary quality control and imputation of the genotype data were described elsewhere. We performed further quality control to filter SNPs with a minor allele frequency of at least 0.01 and a minimum imputation r2 of 0.8 and that were in Hardy–Weinberg equilibrium (HWE; P < 1 × 10−6). After quality control, a total of 7,725,440 SNPs were included in this study. For quality control of participants, we excluded individuals with non-European ancestry, outliers in ancestry and mismatches in sex, resulting in 396,443 participants remaining.

### Polygenetic risk score (PRS)

The base data for the PRS of MDD was based on a multi-ancestry meta-analysis of MDD GWAS results published by Meng et al. in 2024, which included 88,316 MDD cases and 902,757 controls. To avoid sample overlap, we excluded data from the UKB and generated a new GWAS meta-analysis result using the remaining 20 cohorts. Detailed descriptions of the GWAS summary statistics are available elsewhere^33^. The base data for the PRS of GAD in this study was based on a meta-analysis of anxiety disorders GWAS results involving 6 large cohorts comprising 97,383 anxiety cases and 1,169,397 controls^34^. To avoid sample overlap, we excluded data from the UK Biobank and generated a new meta-GWAS using the remaining five cohorts.

This study used genomic data from the UK Biobank as the target data. PRSs were calculated using SNPs passing QC (MAF > 1%). PRSs for both phenotypes were calculated using PRS-CS^35^. After calculating the effect sizes for SNPs, the posterior effect sizes of SNPs were measured using PLINK 2.0, and all SNPs were aggregated into the PRS for each individual.

### Statistical analysis

#### Environmental variables screening

In this study, we utilized the PHESANT package in R (version 4.3) to screen environmental variables associated with three outcomes^36^. For multi-category variables, each category was converted into a binary variable. For variables containing missing value indicators (e.g., negative numbers representing “don’t know” or “prefer not to answer”), the corresponding participants were excluded. 20 questions were included in this study based on a literature review, and 36 variables were transformed after processing. Logistic regression models were used in this package, with gender, age, and assessment centre used as covariates. Standardised regression coefficients (β) were estimated as effect sizes, and all coefficients were converted to odds ratios (OR). All analyses applied two-sided statistical tests. Bonferroni correction was used to control for multiple comparisons (α = 0.05).

#### Gene-environment interaction analyses

We utilized two methods to investigate if gene-environment interactions have effects on the onset of MDD and GAD. Firstly, we constructed a logistic model with PRS, environmental variables and their interaction as independent variables. Then we utilized fastGWA-GE from GCTA to screen for susceptibility to significant environmental variables.

#### PRS×E logistic models

Logistic models were constructed with PRS, screened environmental factors and their interaction (PRS*E) as independent variables, and sex, age, 10 PCs and assessment centres as covariates. This model was used by previous gene-environment interaction studies^37^. The equation is listed below.

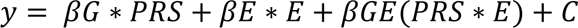

In this equation, y represents the outcome, E represents environmental factor, C is the adjusted covariates, including sex, age, 10 PCs and assessment centres. Bonferroni corrections were utilized to control multiple comparisons.

#### GWEIS

In our study, fastGWA-GE was used. It is a fast and powerful tool for genotype-environment interaction tests in large-scale datasets. The standard LMM was used in the model, and the equation of this model was extended from the one in fastGWA. Annotations were conducted in FUMA^38^. Details were described elsewhere^39^.

#### Two-sample MR

To further investigate the causality between environmental factors and outcomes, we utilised the TwoSampleMR package^40^. Due to the lack of public GWASs for many significant environmental factors, we only investigated the causality between childhood maltreatment and outcomes. To ensure the stability, random-effects models such as IVW, WM, and MR Egger were used. Also, leave-one-out tests and MR-PRESSO analyse were conducted to ensure the robustness.

## Result

### The demographic characteristics of participants

In this study, a total of 396,443 participants were included, with 53.9% being female (n=213,839). The average age of the participants was 57 ± 8 years. In terms of educational attainment, 30.6% of the participants (n = 121,241) had a bachelor or higher degree. In terms of employment status, 56.5% of the participants were employed, while 43.5% were in other statuses, including retired, unemployed, taking care of family, unable to work due to disability or illness, volunteering, and full-time students. The average Townsend Deprivation Index (TDI) was -1.6 ± 3. These results showed that the majority of the participants included in this study were middle-aged individuals with higher educational attainment, higher economic status, and lower levels of deprivation. After grouping participants into the MDD-only group (n = 58,582), GAD-only group (n = 12,063), comorbid group (n = 26,119), and control group (n = 299,679), four groups showed significant differences in the distribution of demographic characteristics (P < 0.001). To better compare the differences in the demographic features of the three groups, pairwise comparisons were conducted. The comorbid group had the highest proportion of female (68.3%), which was significantly higher than the MDD-only group (64.5%) and the GAD-only group (64.2%). The mean age of the GAD-only group was the highest compared to the other two groups. There were significant differences in the distribution of educational attainment, employment status and TDI between the MDD-only group, GAD-only group and comorbid group, while there were no significant differences in the distribution of these variables between the MDD-only group and the comorbid group. The GAD-only group had a lower overall educational level compared to the other two groups. Employment status showed that the MDD-only group (59%) and the comorbid group (57.3%) had significantly higher proportions of employed individuals compared to the GAD-only group (51.5%), while the GAD-only group had a higher proportion of retired individuals (42.7%) compared to the other two groups (Figure 1 and Table S2).

**Figure 1.**
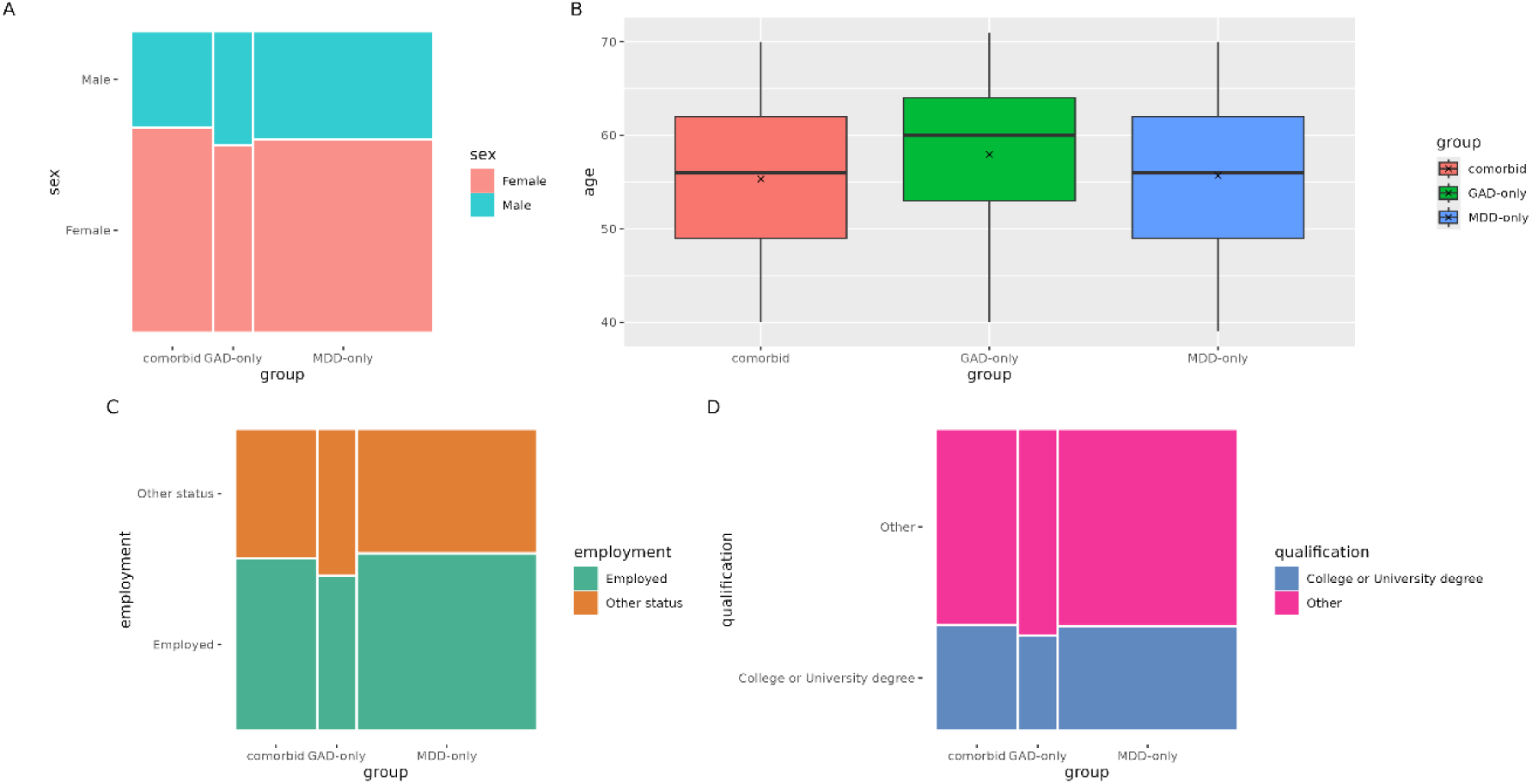
Socio-demographic features of MDD-only, GAD-only and Comorbid group. #Notes: A: Comparison of sex percentages among MDD-only, GAD-only and Comorbid group; B: distribution of age among MDD-only, GAD-only and Comorbid group; C: difference among MDD-only, GAD-only and Comorbid group; D: distribution of age among MDD-only, GAD-only and Comorbid group

### Environmental variables screening results

Using PHESANT, 32,21, and 31 of 36 variables passed the Bonferroni-corrected threshold and were associated with the outcomes of MDD-only, GAD-only and comorbid groups. These variables were included into three dimensions: socio-demographic, early life experiences and lifestyle. 12,7, and 11 of the 14 socio-demographic variables passed Bonferroni correction and were significantly associated with the outcomes of MDD-only, GAD-only, and comorbid group, respectively (Figure 2A). 7,5, and 7 early life experience variables were found to be significantly associated with the three outcomes (Figure 2B). In lifestyle domain, 13,9, and 13 variables were significantly associated with the outcomes significantly lifestyle (Figure 2C). In total, there were 19 significant variables shared among the three groups, including stressful life events, childhood maltreatment, digital screen use, smoking, obesity, substance use, etc (Figure 2D). Taking care of family and TV screen use time were the unique factor for the MDD-only group. Retirement was the unique factors for the GAD-only group. After excluding the comorbid group, there were no shared variables between the MDD-only and GAD-only groups.

**Figure 2.**
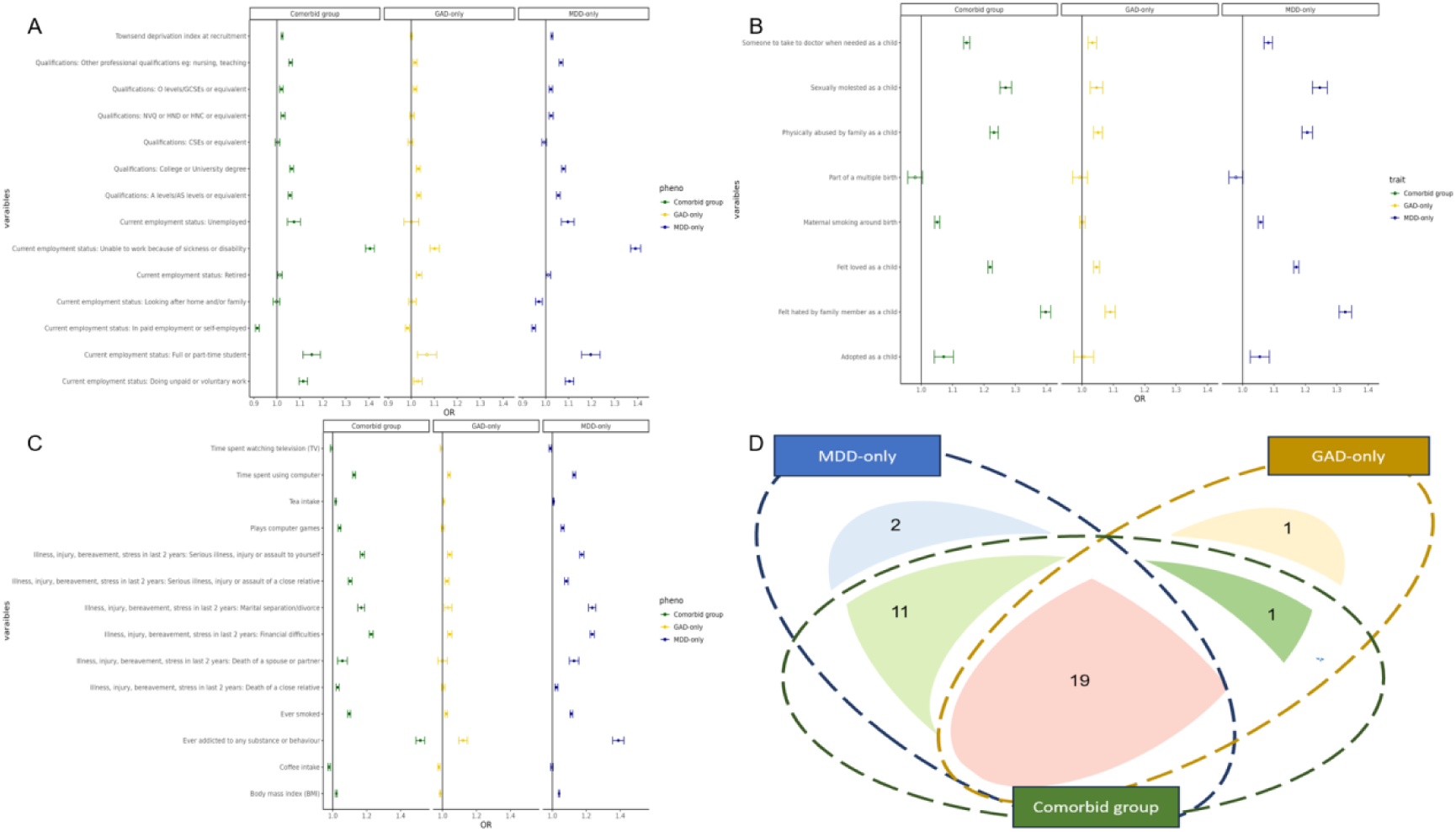
Screened environments from socio-demographic, early life experiences and lifestyle domains. #Notes: A: Comparison of sex percentages among MDD-only, GAD-only and Comorbid group; B: distribution of age among MDD-only, GAD-only and Comorbid group; C: difference among MDD-only, GAD-only and Comorbid group; D: distribution of age among MDD-only, GAD-only and Comorbid group

### Gene-environment interaction results

The effect of the interaction between PRS and environmental factors on three outcomes Across all models, the PRS for MDD and GAD significantly predicted MDD-only, GAD-only and comorbidity (ORs= 1.11-1.15; P values: 9.1×10^-83^ to 1.62×10^-150^). However, among all screened environmental variables, no significant interactions passed Bonferroni correction (Table S2-4).

We analysed the SNP-environment interaction between 32, 21, and 31 environmental factors and 7,725,440 SNPs for three phenotypes, respectively. For MDD-only, 7 significant SNP-environment interactions were found to reach genome-wide significance (P = 5×10^-8^), in which we identified 58, 19, 2246, 1153 and 23 new independent loci passed Bonferroni correction (5×10^-8^/32) for 5 stressful life events including serious illness to yourself or close relatives, death of a spouse, divorce and financial difficulties. (Table 1, Figures S2-8). 8 significant SNP-environment interactions reached genome-wide significance (P = 5×10^-8^) to affect GAD-only. We identified 4,51,1,4,5,4,3, and 3 new independent loci that passed Bonferroni correction (5×10^-8^/21) for 4 childhood maltreatment, time spent on TV and 3 stressful life events (Table 1, Figures S9-16). 7 significant SNP-environment interactions reached genome-wide significance (P = 5×10^-8^), of which 90, 1 and 163 new independent loci were identified to have significant interactions with 3 environments including stressful life events, being adopted and time spent on computers affecting comorbidity (5×10^-8^/31) (Table 1, Figures S17-23).

**Table 1.**
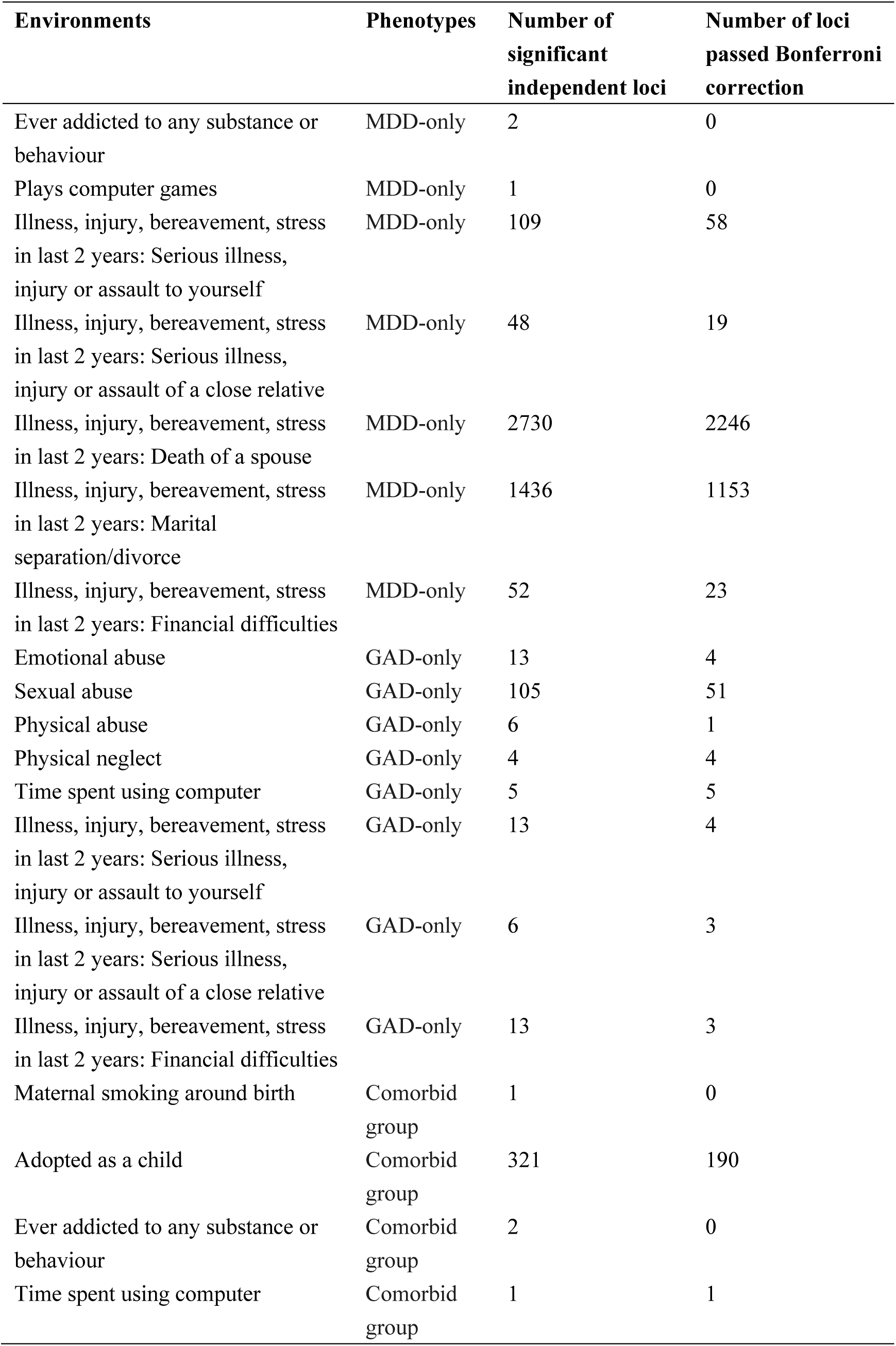
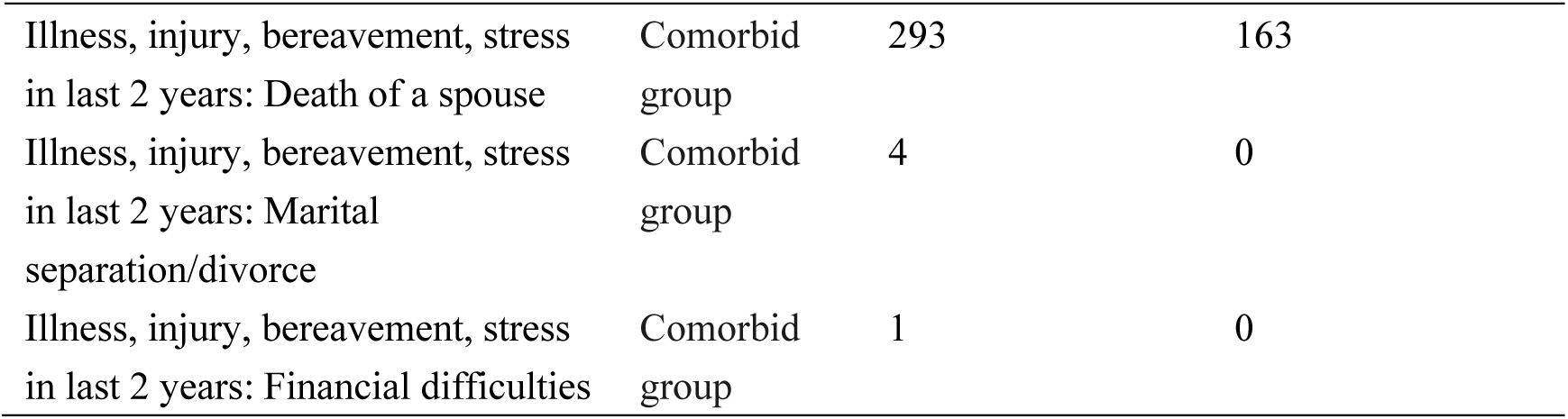
SNP-environment interaction results in MDD-only, GAD-only and Comorbid group.

To interpret of the biology function of GWEIS findings, we then implemented gene-based tests for 19310 protein-coding genes using MAGMA, to investigate whether variation in particular genes tend to modulate the impact of environmental exposures on phenotypes. For MDD-only, we found 64 genes in total from 5 environments reached the standard genome-wide significance (P< 2.59×10^-6^, 0.05/19310), of which 41 genes survived Bonferroni correction (2.59×10^-6^/32); one of two genes that reached genome-wide significance passed Bonferroni correction for one environment in GAD-only; all of the three genes for three environments passed Bonferroni correction for comorbid MDD/GAD (Table 2).

**Table 2.** Gene-environment interaction results in MDD-only, GAD-only and Comorbid group.

| **Environments** | **Phenotypes** | **Chromosome** | **Genes** | **P** |
| --- | --- | --- | --- | --- |
| Illness, injury, bereavement, stress<br>in last 2 years: Serious illness,<br>injury or assault to yourself | MDD-only | 18 | RIT2 | 1.05E-06 |
| Illness, injury, bereavement, stress<br>in last 2 years: Serious illness,<br>injury or assault of a close relative | MDD-only | 6 | RAB23 | 1.98E-06 |
| Illness, injury, bereavement, stress<br>in last 2 years: Death of a close<br>relative | MDD-only | 1 | NOTCH2NL | 1.94E-15 |
|  | MDD-only | 1 | RP11-458D21.5 | 1.84E-12 |
|  | MDD-only | 1 | FCGR3B | 1.37E-08 |
|  | MDD-only | 1 | CRB1 | 1.33E-08 |
|  | MDD-only | 1 | OR2T10 | 7.44E-07 |
|  | MDD-only | 2 | CFC1B | 1.35E-13 |
|  | MDD-only | 3 | IL5RA | 1.76E-11 |
|  | MDD-only | 3 | TRNT1 | 4.93E-11 |
|  | MDD-only | 3 | CRBN | 2.93E-07 |
|  | MDD-only | 3 | FRMD4B | 5.62E-09 |
|  | MDD-only | 3 | ADCY5 | 1.98E-06 |
|  | MDD-only | 3 | ETV5 | 1.79E-07 |
|  | MDD-only | 4 | KIAA1109 | 5.21E-07 |
|  | MDD-only | 4 | ADAD1 | 1.14E-07 |
|  | MDD-only | 4 | SLC7A11 | 2.25E-09 |
|  | MDD-only | 7 | UNCX | 6.64E-07 |
|  | MDD-only | 7 | ARMC10 | 4.04E-11 |
|  | MDD-only | 7 | NAPEPLD | 1.51E-08 |
|  | MDD-only | 8 | LONRF1 | 2.04E-08 |
|  | MDD-only | 9 | SPATA31A3 | 1.55E-15 |
|  | MDD-only | 9 | TNC | 8.93E-07 |
|  | MDD-only | 10 | SYT15 | 2.81E-08 |
|  | MDD-only | 10 | GPRIN2 | 3.61E-09 |
|  | MDD-only | 10 | ANTXRL | 8.91E-08 |
|  | MDD-only | 11 | CASP12 | 3.15E-07 |
|  | MDD-only | 12 | ETV6 | 6.29E-07 |
|  | MDD-only | 13 | SERTM1 | 4.30E-10 |
|  | MDD-only | 14 | PACS2 | 4.12E-07 |
|  | MDD-only | 14 | TEX22 | 1.00E-09 |
|  | MDD-only | 15 | TMEM87A | 1.11E-08 |
|  | MDD-only | 15 | GANC | 6.69E-08 |
|  | MDD-only | 15 | TLN2 | 5.93E-08 |
|  | MDD-only | 15 | RP11-625H11.1 | 2.72E-10 |
|  | MDD-only | 16 | TP53TG3 | 1.89E-13 |
|  | MDD-only | 17 | RPH3AL | 1.06E-06 |
|  | MDD-only | 17 | ICAM2 | 2.18E-06 |
| Illness, injury, bereavement, stress<br>in last 2 years: Marital<br>separation/divorce | MDD-only | 1 | RHD | 4.91E-07 |
|  | MDD-only | 1 | NOTCH2NL | 1.04E-10 |
|  | MDD-only | 1 | RP11-458D21.5 | 9.04E-08 |
|  | MDD-only | 1 | FCGR3B | 1.88E-07 |
|  | MDD-only | 1 | GUK1 | 7.47E-10 |
|  | MDD-only | 1 | GJC2 | 2.32E-09 |
|  | MDD-only | 1 | IBA57-AS1 | 1.79E-08 |
|  | MDD-only | 4 | UGT2B17 | 7.31E-07 |
|  | MDD-only | 4 | CLCN3 | 1.75E-07 |
|  | MDD-only | 9 | FOXD4L6 | 5.00E-10 |
|  | MDD-only | 9 | CBWD6 | 5.00E-10 |
|  | MDD-only | 9 | BX255923.1 | 5.00E-10 |
|  | MDD-only | 10 | ECHDC3 | 6.92E-09 |
|  | MDD-only | 10 | NPY4R | 5.67E-08 |
|  | MDD-only | 10 | ANXA8L1 | 3.14E-07 |
|  | MDD-only | 13 | NUPL1 | 1.07E-08 |
|  | MDD-only | 13 | FLT3 | 2.43E-06 |
|  | MDD-only | 15 | CHRFAM7A | 2.43E-09 |
|  | MDD-only | 15 | GOLGA8R | 7.65E-11 |
|  | MDD-only | 16 | TP53TG3 | 7.81E-14 |
|  | MDD-only | 17 | C17orf58 | 1.51E-06 |
| Illness, injury, bereavement, stress<br>in last 2 years: Financial<br>difficulties | GAD-only | 1 | NVL | 1.24E-07 |
|  | GAD-only | 13 | DLEU1 | 7.64E-07 |
| Adopted as a child | Comorbid<br>group | 3 | HESX1 | 2.35E-08 |
| Illness, injury, bereavement, stress<br>in last 2 years: Death of a spouse or<br>partner | Comorbid<br>group | 13 | CDK8 | 8.98E-10 |
| Illness, injury, bereavement, stress<br>in last 2 years: Financial<br>difficulties | Comorbid<br>group | 2 | PDE1A | 3.12E-07 |

Across all significant SNPs-environment interactions that reached genome-wide significance, SNPs-financial difficulties interaction was common among MDD-only, GAD-only and comorbid MDD/GAD, however, the specific loci showing significant interactions with financial difficulties were distinct across the phenotypes (Figure 3). No common genes that significantly interact with environments were found across phenotypes.

**Figure 3.**
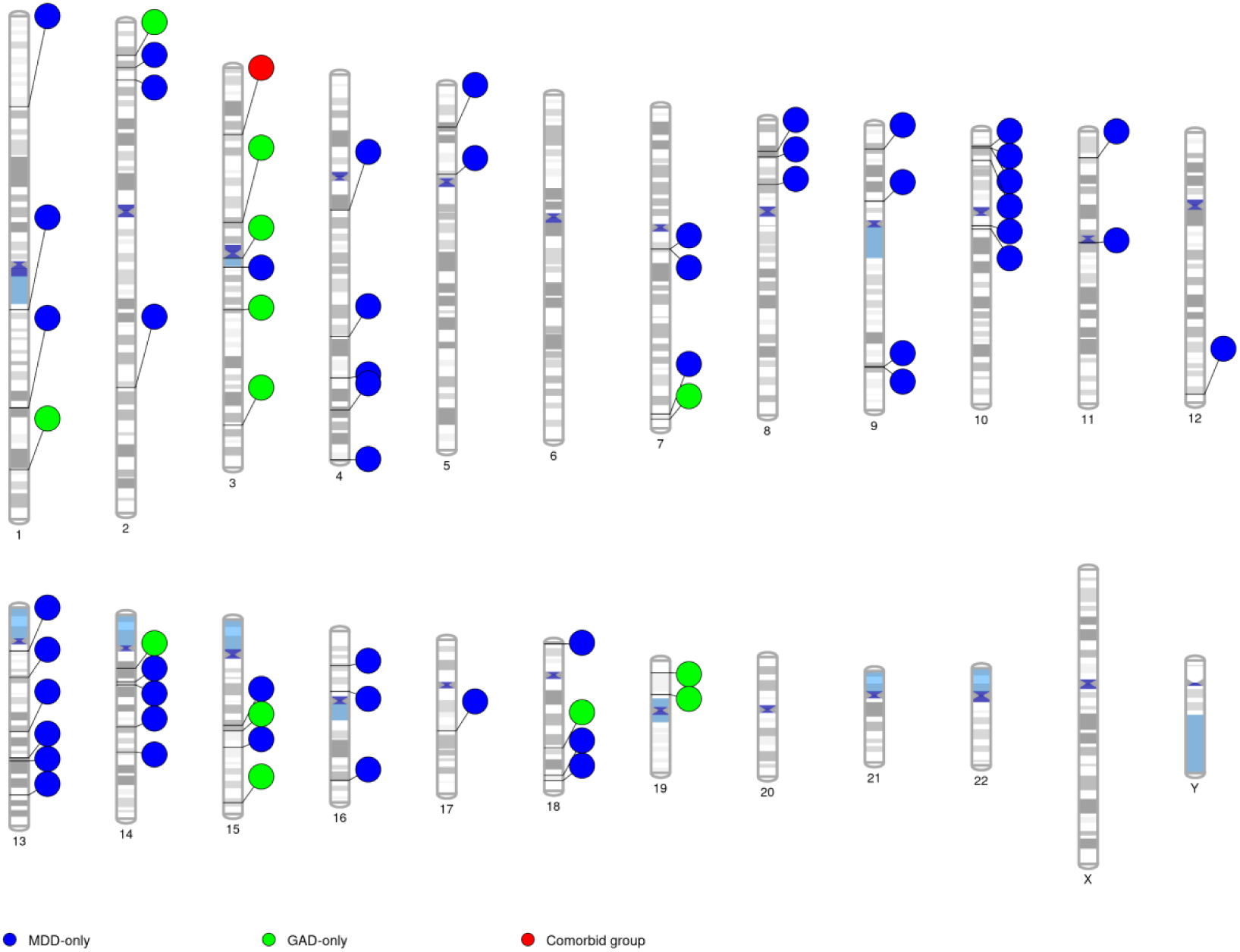
Location of SNPs interacting with financial difficulties for three phenotypes. #Notes: An Ideogram of all chromosomes is plotted. Lines are plotted on the chromosomes corresponding to the base-pair location of each SNP, and the line connects to coloured circles representing the phenotypes associated with that SNP. Blue, green and red circles represent MDD-only, GAD-only and comorbid group phenotypes, respectively.

Gene childhood maltreatment interaction was only found to significantly affect the onset of GAD-only. To further investigate whether childhood maltreatment was heritable, we utilized GWAS summary data from other database to analysed genetic correlations with three phenotypes. Results showed that moderate genetic correlations between childhood maltreatment and MDD-only (rg = 0.55, SE = 0.035, P = 1.53×10⁻⁵⁵) and comorbid MDD/GAD (rg = 0.56, SE = 0.036, P = 2.97 ×10⁻⁵⁴). Low genetic correlation (rg = 0.25, SE = 0.061, P = 5.40×10⁻⁵) was found between GAD-only and childhood maltreatment.

To further understand the relationship between childhood maltreatment and phenotypes, two sample MR was utilized to investigate the causality between them. Forward MR showed that childhood maltreatment causally increases the risk of MDD-only, GAD-only and their comorbidity. The inverse-variance weighted (IVW) estimate supported a positive effect of childhood maltreatment on the of risk MDD-only (OR_IVW = 1.97, P_IVW = 1.94×10⁻⁷) (Figure 4A and Table S5); the weighted-median (WM) and MR-Egger estimates were directionally consistent with IVW. For GAD-only, IVW likewise indicated an increased risk (OR_IVW = 2.24, P_IVW = 0.0039) (Figure 4B and Table S5), with WM concordant in direction; although the MR-Egger point estimate was in the opposite direction, the MR-Egger intercept was not statistically significant (P_intercept = 0.067), providing no evidence of directional pleiotropy and supporting the reliability of the IVW result. Both IVW and WM analyses supported a causal effect of childhood maltreatment on comorbid MDD/GAD (OR_IVW = 3.06, P_IVW = 5.28×10⁻⁷; OR_WM = 2.69, P_WM = 5.10×10⁻⁶) ((Figure 4C and Table S5). Although MR-Egger yielded an effect estimate in the opposite direction, sensitivity analyses using MR-PRESSO did not detect horizontal pleiotropy or outlier SNPs, further corroborating the validity of the IVW findings. Reverse MR analyses provided no evidence for a causal effect of MDD-only, GAD-only, or their comorbidity on childhood maltreatment.

**Figure 4.**
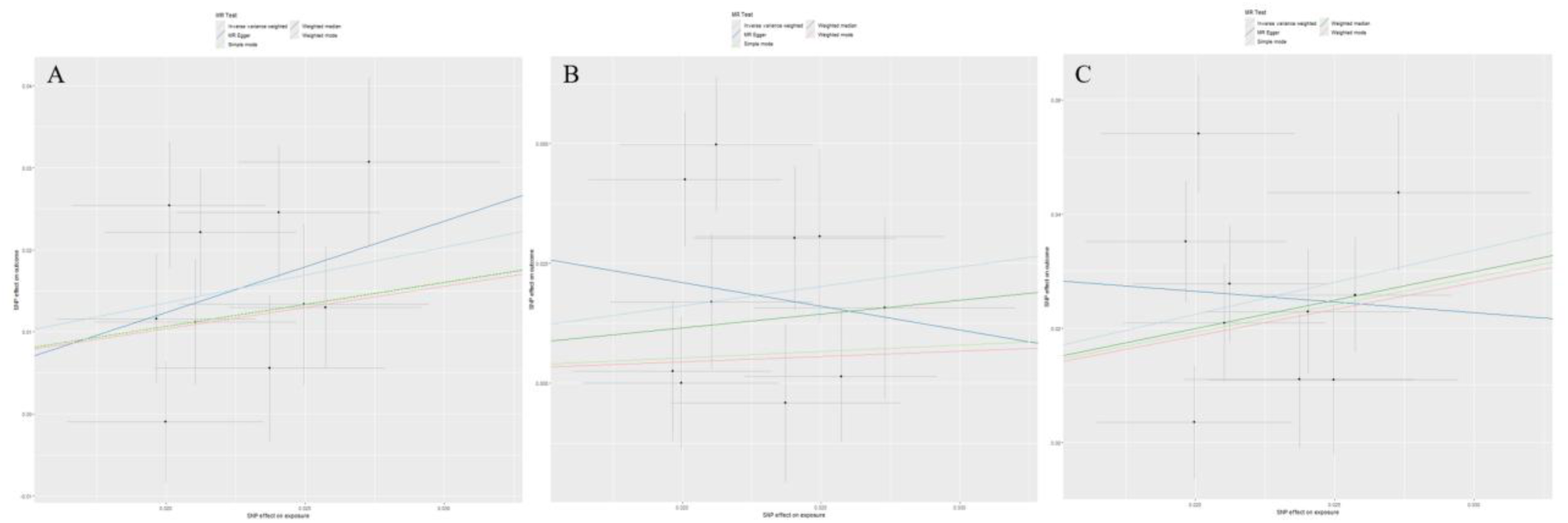
Two-sample MR analyses results for causal relationship between childhood maltreatment and three phenotypes. #Notes: The horizontal axis represents the effect size of the SNP on exposure, while the vertical axis denotes the effect size of the SNP on the outcome. The differently coloured lines above indicate results obtained from various MR methods; A: Two-sample MR results from childhood adversity to MDD-only; B: Two-sample MR results from childhood adversity to GAD-only; C: Two-sample MR results from childhood adversity to comorbidity.

## Discussion

To our knowledge, this is the first study to systematically compare socio-demographic features, associated environmental factors and gene-environment interaction patterns of MDD and GAD, dividing them into three case groups (MDD-only, GAD-only and their comorbid group). Across 36 environmental variables, we identified partially overlapping environments in the three groups. Genome-wide gene-environment interaction analyses revealed novel loci exhibiting significant gene–environment (G×E) interactions across all groups, conferring elevated liability to disorder onset. In the MDD-only group, risk loci demonstrated robust interactions with five environmental exposures, including recent life stressors. Conversely, loci associated with GAD-only exhibited significant interactions with eight exposures, including childhood maltreatment and recent stress. Only three environmental variables passed Bonferroni correction - including recent stress and adoption history-interacted with genetic variants to amplify comorbidity risk. Importantly, the three groups displayed different G×E interaction patterns. While all groups exhibited significant SNP–financial difficulty interactions, the specific susceptibility loci implicated in these interactions were distinct across groups.

Significant difference in socio-demographic features across groups was found, indicating that individuals with clinically worse MDD (MDD comorbid with GAD) were more likely to be women with lower socioeconomic status, whereas males with pure GAD were more likely to have lower educational attainment and retired status, which is consistent with previous epidemiological investigations. Our findings confirm that economic stress, childhood maltreatment, stressful life events, and unhealthy lifestyle behaviours constitute shared environmental risk factors for both MDD and GAD^6,41,42^. We also identified maternal smoking during pregnancy as a unique environmental risk factor for MDD and comorbid MDD/GAD, consistent with prior evidence showing no significant association between maternal smoking and offspring’s an increased risk for depression^43^. This finding underscores the influence of prenatal exposures on neurodevelopment^44^. Conversely, coffee consumption frequency emerged as a unique protective factor for GAD. However, higher coffee intake was thought to exacerbates anxiety symptoms^45^. Recent evidence suggests a J-shaped association between coffee consumption and anxiety; whereby moderate intake may confer protective effects^46^. In our sample, the average coffee consumption frequency was relatively low, which may account for its protective association.

We found that the PRS for both MDD and GAD significantly increased susceptibility to these disorders. However, no significant interactions were observed between any PRS and significant environmental factors in influencing the risk of MDD or GAD. Previous results for PRS-environment interactions were inconsistent. Peyrot et al. conducted a meta-analysis across nine cohorts and found no significant interactions between MDD PRS and five dimensions of childhood maltreatment^37^. Conversely, other studies have reported significant interactions between MDD PRS and childhood maltreatment but with inconsistent direction of effect sizes for these interaction terms, suggesting limited statistical power for detecting interactions^47-49^. Therefore, the null findings in the present study could be related to insufficient power of the interaction models. Rujia et al. reported that stress exposure significantly modified the effect of PRS on depression and anxiety^50^. Although this finding differs from ours, it provides a plausible explanation for our null results. Rujia et al. calculated PRS using SNP weights across multiple p-value thresholds and found significant interactions between PRS and stress when SNPs with p-values between 0.1 and 0.05 were included. In contrast, our study employed PRS-CS, which adaptively shrinks SNP effect sizes based on their p-values, assigning greater weight to genome-wide significant SNPs. Consequently, our findings may primarily reflect insignificant interactions between main-effect variants and environmental exposures. These results underscore the importance of genome-wide gene–environment interaction analyses, which can identify variants with G×E effects that do not reach genome-wide significance in GWAS.

Using fastGWA-GE, we conducted genome-wide G×E analyses and identified distinct interaction patterns across MDD-only, GAD-only, and comorbid MDD/GAD. Notably, we are the first to report significant interactions between genetic variants and environmental factors such as substance use and adoption history in MDD and GAD. Previous studies have primarily focused on interactions between genetic variants and stress/childhood maltreatment, our findings partly alien with them. We found that stressful life events interact with distinct genetic loci to influence MDD and GAD. To date, only two large-scale GWEIS have examined gene-stress interactions in depression: Arnau-Soler et al. identified two significant SNPs near PIWIL4 and within ZCCHC2 using Generation Scotland^23^, while Suppli et al. reported three significant SNPs in ABCC1, AKAP6, and MSFD1^51^. Our results replicated part of their findings. The significant SNPs interacted with stress in MDD-only can be mapped to four of these genes (ABCC1, AKAP6, PIWIL4, ZCCHC2), with two of which mapped to ZCCHC2 and AKAP6 in MDD/GAD comorbid group. Meta-analytic evidence links AKAP6 to general cognitive function^52^, and PIWIL4 to neurocognitive processes^53^. SNP within the AKAP6 gene were found to have a negative correlation with stress, with increased stress associated with a reduced risk of depression^51^. A review indicated that higher cognitive abilities may serve as a protective factor, lowering the risk of developing depression^54^. Findings from this study demonstrate that the interaction between the AKAP6 and PIWIL4 genes and stress enhances individuals’ adaptability to high-pressure environments by elevating cognitive function, thereby reducing the risk of depression onset. This aligns with theories of differential susceptibility and plasticity^55^, which posit that certain genotypes confer resilience under adverse environments. ABCC1 encodes MRP1, a transporter expressed at the blood–brain barrier, while ZCCHC2 encodes a zinc-finger protein expressed in CD4+ T cells. Variants in these genes have been linked to antidepressant response, particularly for citalopram^56,57^. Both of the prior UKB-based studies failed to replicate these findings. We kept six stress-related events individually, consistent with Generation Scotland and iPSYCH2012 original methodologies. This approach likely enhanced reproducibility and underscores the importance of harmonized environmental measures^58^. Overall, our findings indicate that stress interacts with genetic factors to substantially increase MDD and GAD risk. Notably, most significant SNPs identified in our GWEIS did not reach genome-wide significance in standard GWAS, highlighting the value of G×E analyses for uncovering biologically meaningful variants missed by main-effect models.

Significant gene and childhood maltreatment interactions for GAD-only were found, whereas no significant G×E effects were detected for MDD-only or comorbid group. This partially aligns with recent findings, with significant interacting with childhood maltreatment total scores but not with any dimension^59^. Gene–environment correlation may complicate interpretation, in which genetic factors can influence environmental exposure^60^. Strong genetic correlations among childhood maltreatment, MDD-only and comorbid group were observed, but correlations with GAD-only were weaker. Previous findings confirmed that the gene-environment correlation can be mixed up with G×E. Varun et al. identified a gene–environment correlation between childhood adversities and depression, and a unidirectional causal pathway from adversities to depression using Mendelian Randomization analyses, which is consistent with our findings^61^. We are the first to examine genome-wide gene and childhood maltreatment interactions in GAD. Although we identified significant interactions, the implicated genes did not overlap with previously reported candidates. These findings suggest that while childhood maltreatment is a shared risk factor for MDD and GAD, the underlying G×E mechanisms may differ across disorders.

Finally, we wish to point several limitations of this study. First, as a retrospective study, recall bias is inevitable. Second, causal relationships between some certain environments and the disorders are limited by public GWAS summary statistics. Lastly, we lack replications to another independent cohort. However, we were able to replicate some findings from independent cohorts reported in previous studies, which partially mitigates this limitation.

## Conclusions

By comparing the MDD-only, GAD-only, and comorbid groups on socio-demographic, environmental, and gene-environment interaction dimensions, we found that there were partially overlapping environmental factors among the three groups, partially shared genes between the depression and comorbid groups, and different patterns of gene-environment interactions among the three groups. This finding preliminarily confirms the existence of relatively specific environmental factors and the gene-interaction patterns between MDD and GAD, which may be potential biological evidence for the different clinical features of the two diseases. This work provides preliminary evidence for unique G×E mechanisms in MDD, GAD and their comorbidity, offering a foundation for future subtype-focused research and targeted interventions. Further research is needed to explore the underlying mechanisms and to identify additional variables that may contribute to these differences.

## Acknowledgement

The authors would like to thank all contributions who were involved in establishing the UK Biobank and extend our appreciation to all participants who generously shared their information.

## Data availability

Data is available on the UK Biobank website: https://www.ukbiobank.ac.uk/.

## References

1. Huang Y, Wang Y, Wang H, et al. Prevalence of mental disorders in China: a cross-sectional epidemiological study. The Lancet Psychiatry. 2019;6(3):211–224. doi:10.1016/s2215-0366(18)30511-x

2. Hammen C. Risk factors for depression: an autobiographical review. Annual review of clinical psychology. 2018;14(1):1–28.

3. Lund C, Breen A, Flisher AJ, et al. Poverty and common mental disorders in low and middle income countries: A systematic review. Social science & medicine. 2010;71(3):517–528.

4. Hoppen TH, Chalder T. Childhood adversity as a transdiagnostic risk factor for affective disorders in adulthood: A systematic review focusing on biopsychosocial moderating and mediating variables. Clinical Psychology Review. Nov 2018;65:81–151. doi:10.1016/j.cpr.2018.08.002

5. Wang R, Shi M, Zhang Q, et al. The association of early life factors with depression and anxiety in adults aged 40–69 years: a population-based cohort study. Translational Psychiatry. 2024/07/20 2024;14(1):299. doi:10.1038/s41398-024-03006-7

6. Firth J, Siddiqi N, Koyanagi A, et al. The Lancet Psychiatry Commission: a blueprint for protecting physical health in people with mental illness. The lancet psychiatry. 2019;6(8):675–712.

7. Stubbs B, Koyanagi A, Hallgren M, et al. Physical activity and anxiety: A perspective from the World Health Survey. Journal of affective disorders. 2017;208:545–552.

8. Virtanen S, Kuja-Halkola R, Mataix-Cols D, et al. Comorbidity of substance misuse with anxiety-related and depressive disorders: a genetically informative population study of 3 million individuals in Sweden. Article. Psychological Medicine. Jul 2020;50(10):1706–1715. Pii s0033291719001788. doi:10.1017/s0033291719001788

9. Stubbs B, Vancampfort D, Firth J, et al. Association between depression and smoking: a global perspective from 48 low-and middle-income countries. Journal of psychiatric research. 2018;103:142–149.

10. Gonda X, Hullam G, Antal P, et al. Significance of risk polymorphisms for depression depends on stress exposure. Sci Rep. Mar 2 2018;8(1):3946. doi:10.1038/s41598-018-22221-z

11. Roberston L, Twenge JM, Joiner TE, Cummins K. Associations between screen time and internalizing disorder diagnoses among 9- to 10-year-olds. Journal of Affective Disorders. 2022/08/15/ 2022;311:530-537. 10.1016/j.jad.2022.05.071

12. Sullivan PF, Neale MC, Kendler KS. Genetic epidemiology of major depression: review and meta-analysis. American journal of psychiatry. 2000;157(10):1552–1562.

13. Newman SC, Bland RC. A population-based family study of DSM-III generalized anxiety disorder. Psychological medicine. 2006;36(9):1275–1281.

14. Caspi A, Sugden K, Moffitt TE, et al. Influence of life stress on depression: moderation by a polymorphism in the 5-HTT gene. Science. 2003;301(5631):386–389.

15. Thorp JG, Campos AI, Grotzinger AD, et al. Symptom-level modelling unravels the shared genetic architecture of anxiety and depression. Nat Hum Behav. Oct 2021;5(10):1432–1442. doi:10.1038/s41562-021-01094-9

16. Purves KL, Coleman JRI, Meier SM, et al. A major role for common genetic variation in anxiety disorders. Mol Psychiatry. Dec 2020;25(12):3292–3303. doi:10.1038/s41380-019-0559-1

17. Kendler KS, Neale MC, Kessler RC, Heath AC, Eaves LJ. Major depression and generalized anxiety disorder. Same genes, (partly) different environments? Arch Gen Psychiatry. Sep 1992;49(9):716–22. doi:10.1001/archpsyc.1992.01820090044008

18. Tao Y, Zhao R, Yang B, Han J, Li Y. Dissecting the shared genetic landscape of anxiety, depression, and schizophrenia. Journal of Translational Medicine. 2024/04/18 2024;22(1):373. doi:10.1186/s12967-024-05153-3

19. Uher R, Zwicker A. Etiology in psychiatry: embracing the reality of poly-gene-environmental causation of mental illness. World Psychiatry. Jun 2017;16(2):121–129. doi:10.1002/wps.20436

20. Kim-Cohen J, Caspi A, Taylor A, et al. MAOA, maltreatment, and gene-environment interaction predicting children’s mental health: new evidence and a meta-analysis. Mol Psychiatry. Oct 2006;11(10):903–13. doi:10.1038/sj.mp.4001851

21. Dunn EC, Wiste A, Radmanesh F, et al. GENOME-WIDE ASSOCIATION STUDY (GWAS) AND GENOME-WIDE BY ENVIRONMENT INTERACTION STUDY (GWEIS) OF DEPRESSIVE SYMPTOMS IN AFRICAN AMERICAN AND HISPANIC/LATINA WOMEN. Depression and Anxiety. 2016;33(4):265–280. 10.1002/da.22484

22. Werme J, van der Sluis S, Posthuma D, de Leeuw CA. Genome-wide gene-environment interactions in neuroticism: an exploratory study across 25 environments. Translational Psychiatry. 2021;11(1)doi:10.1038/s41398-021-01288-9

23. Arnau-Soler A, Macdonald-Dunlop E, Adams MJ, et al. Genome-wide by environment interaction studies of depressive symptoms and psychosocial stress in UK Biobank and Generation Scotland. Transl Psychiatry. Feb 4 2019;9(1):14. doi:10.1038/s41398-018-0360-y

24. Flint J. The genetic basis of major depressive disorder. Molecular Psychiatry. 2023/06/01 2023;28(6):2254-2265. doi:10.1038/s41380-023-01957-9

25. Nguyen TD, Harder A, Xiong Y, et al. Genetic heterogeneity and subtypes of major depression. Mol Psychiatry. Mar 2022;27(3):1667–1675. doi:10.1038/s41380-021-01413-6

26. Lamers F, van Oppen P, Comijs HC, et al. Comorbidity patterns of anxiety and depressive disorders in a large cohort study: the Netherlands Study of Depression and Anxiety (NESDA). J Clin Psychiatry. Mar 2011;72(3):341–8. doi:10.4088/JCP.10m06176blu

27. Carter RM, Wittchen HU, Pfister H, Kessler RC. One-year prevalence of subthreshold and threshold DSM-IV generalized anxiety disorder in a nationally representative sample. Depression and anxiety. 2001;13(2):78–88.

28. van Loo HM, Schoevers RA, Kendler KS, de Jonge P, Romeijn JW. Psychiatric comorbidity does not only depend on diagnostic thresholds: an illustration with major depressive disorder and generalized anxiety disorder. Depression and anxiety. 2016;33(2):143–152.

29. Howard DM, Adams MJ, Shirali M, et al. Genome-wide association study of depression phenotypes in UK Biobank identifies variants in excitatory synaptic pathways. Nature Communications. 2018/04/16 2018;9(1):1470. doi:10.1038/s41467-018-03819-3

30. Bycroft C, Freeman C, Petkova D, et al. The UK Biobank resource with deep phenotyping and genomic data. Nature. 2018/10/01 2018;562(7726):203-209. doi:10.1038/s41586-018-0579-z

31. Cai N, Revez JA, Adams MJ, et al. Minimal phenotyping yields genome-wide association signals of low specificity for major depression. Nature Genetics. 2020/04/01 2020;52(4):437-447. doi:10.1038/s41588-020-0594-5

32. Davis KAS, Cullen B, Adams M, et al. Indicators of mental disorders in UK Biobank-A comparison of approaches. Int J Methods Psychiatr Res. Sep 2019;28(3):e1796. doi:10.1002/mpr.1796

33. Meng X, Navoly G, Giannakopoulou O, et al. Multi-ancestry genome-wide association study of major depression aids locus discovery, fine mapping, gene prioritization and causal inference. Nat Genet. Feb 2024;56(2):222–233. doi:10.1038/s41588-023-01596-4

34. Friligkou E, Løkhammer S, Cabrera-Mendoza B, et al. Gene discovery and biological insights into anxiety disorders from a large-scale multi-ancestry genome-wide association study. Nat Genet. Oct 2024;56(10):2036–2045. doi:10.1038/s41588-024-01908-2

35. Ge T, Chen C-Y, Ni Y, Feng Y-CA, Smoller JW. Polygenic prediction via Bayesian regression and continuous shrinkage priors. Nature Communications. 2019/04/16 2019;10(1):1776. doi:10.1038/s41467-019-09718-5

36. Millard LAC, Davies NM, Gaunt TR, Davey Smith G, Tilling K. Software Application Profile: PHESANT: a tool for performing automated phenome scans in UK Biobank. Int J Epidemiol. Feb 2018;47(1):29–35. doi:10.1093/ije/dyx204

37. Peyrot WJ, Van der Auwera S, Milaneschi Y, et al. Does childhood trauma moderate polygenic risk for depression? A meta-analysis of 5765 subjects from the psychiatric genomics consortium. Biological psychiatry. 2018;84(2):138-147.

38. Watanabe K, Umićević Mirkov M, de Leeuw CA, van den Heuvel MP, Posthuma D. Genetic mapping of cell type specificity for complex traits. Nature Communications. 2019/07/19 2019;10(1):3222. doi:10.1038/s41467-019-11181-1

39. Zhong W, Chhibber A, Luo L, Mehrotra DV, Shen J. A fast and powerful linear mixed model approach for genotype-environment interaction tests in large-scale GWAS. Briefings in Bioinformatics. 2022;24(1)doi:10.1093/bib/bbac547

40. Hartwig FP, Davies NM, Hemani G, Davey Smith G. Two-sample Mendelian randomization: avoiding the downsides of a powerful, widely applicable but potentially fallible technique. Int J Epidemiol. Dec 1 2016;45(6):1717–1726. doi:10.1093/ije/dyx028

41. Jaffee SR. Child Maltreatment and Risk for Psychopathology in Childhood and Adulthood. Annu Rev Clin Psychol. May 8 2017;13:525–551. doi:10.1146/annurev-clinpsy-032816-045005

42. Scott KM, Al-Hamzawi AO, Andrade LH, et al. Associations Between Subjective Social Status and DSM-IV Mental Disorders: Results From the World Mental Health Surveys. JAMA Psychiatry. 2014;71(12):1400–1408. doi:10.1001/jamapsychiatry.2014.1337

43. Jareebi MA, Alqassim AY. The impact of educational attainment on mental health: A Causal Assessment from the UKB and FinnGen Cohorts. Medicine. 2024;103(26):e38602. doi:10.1097/md.0000000000038602

44. Chu X, Ye J, Wen Y, et al. Maternal smoking during pregnancy and risks to depression and anxiety in offspring: An observational study and genome-wide gene-environment interaction analysis in UK biobank cohort. Journal of Psychiatric Research. 2021/08/01/ 2021;140:149-158. 10.1016/j.jpsychires.2021.05.067

45. Grosso G, Micek A, Castellano S, Pajak A, Galvano F. Coffee, tea, caffeine and risk of depression: A systematic review and dose–response meta-analysis of observational studies. Molecular Nutrition & Food Research. 2016;60(1):223–234. 10.1002/mnfr.201500620

46. Min J, Cao Z, Cui L, et al. The association between coffee consumption and risk of incident depression and anxiety: Exploring the benefits of moderate intake. Psychiatry Research. 2023/08/01/ 2023;326:115307. 10.1016/j.psychres.2023.115307

47. Chen P, Li Y, Zadrozny S, Seifer R, Belger A. Polygenic risk, childhood abuse and gene x environment interactions with depression development from middle to late adulthood: A U.S. national life-course study. Preventive Medicine. 2024/08/01/ 2024;185:108048. 10.1016/j.ypmed.2024.108048

48. Mullins N, Power R, Fisher H, et al. Polygenic interactions with environmental adversity in the aetiology of major depressive disorder. Psychological medicine. 2016;46(4):759–770.

49. Knol MJ, van der Tweel I, Grobbee DE, Numans ME, Geerlings MI. Estimating interaction on an additive scale between continuous determinants in a logistic regression model. International journal of epidemiology. 2007;36(5):1111–1118.

50. Wang R, Hartman CA, Snieder H, Lifelines Cohort S. Stress-related exposures amplify the effects of genetic susceptibility on depression and anxiety. Translational Psychiatry. 2023/01/30 2023;13(1):27. doi:10.1038/s41398-023-02327-3

51. Suppli NP, Andersen KK, Agerbo E, et al. Genome-wide by Environment Interaction Study of Stressful Life Events and Hospital-Treated Depression in the iPSYCH2012 Sample. Biological Psychiatry Global Open Science. 2022/10/01/ 2022;2(4):400-410. 10.1016/j.bpsgos.2021.11.003

52. Davies G, Armstrong N, Bis JC, et al. Genetic contributions to variation in general cognitive function: a meta-analysis of genome-wide association studies in the CHARGE consortium (N= 53 949). Molecular psychiatry. 2015;20(2):183–192.

53. Rajasethupathy P, Antonov I, Sheridan R, et al. A role for neuronal piRNAs in the epigenetic control of memory-related synaptic plasticity. Cell. 2012;149(3):693–707.

54. Jokela M. Why is cognitive ability associated with psychological distress and wellbeing? Exploring psychological, biological, and social mechanisms. Personality and Individual Differences. 2022/07/01/ 2022;192:111592. 10.1016/j.paid.2022.111592

55. Caspi A, Moffitt TE. Gene–environment interactions in psychiatry: joining forces with neuroscience. Nature Reviews Neuroscience. 2006/07/01 2006;7(7):583-590. doi:10.1038/nrn1925

56. Lee SH, Lee M-S, Lee JH, et al. MRP1 Polymorphisms Associated With Citalopram Response in Patients With Major Depression. Journal of Clinical Psychopharmacology. 2010;30(2)

57. Mamdani F, Berlim MT, Beaulieu M-M, Turecki G. Pharmacogenomic predictors of citalopram treatment outcome in major depressive disorder. The World Journal of Biological Psychiatry. 2014;15(2):135-144.

58. Van der Auwera S, Peyrot WJ, Milaneschi Y, et al. Genome-wide gene-environment interaction in depression: A systematic evaluation of candidate genes: The childhood trauma working-group of PGC-MDD. Am J Med Genet B Neuropsychiatr Genet. Jan 2018;177(1):40–49. doi:10.1002/ajmg.b.32593

59. Sun Y, Liao Y, Zhang Y, et al. Genome-wide interaction association analysis identifies interactive effects of childhood maltreatment and kynurenine pathway on depression. Nature Communications. 2025/02/18 2025;16(1):1748. doi:10.1038/s41467-025-57066-4

60. Rutter M, Thapar A, Pickles A. Gene-environment interactions: biologically valid pathway or artifact? Archives of general psychiatry. 2009;66(12):1287–1289.

61. Warrier V, Kwong ASF, Luo M, et al. Gene-environment correlations and causal effects of childhood maltreatment on physical and mental health: a genetically informed approach. The Lancet Psychiatry. 2021;8(5):373–386. doi:10.1016/S2215-0366(20)30569-1

