## supplement figures 1-23 for "The different gene-environment interaction pattern in major depressive disorder and generalised anxiety disorder: a comparative study across 36 environments"

**table of content**

Figure S1: Flowchart	3

GWEIS Plots for MDD-only

Figure S2: GWEIS Manhattan plot of SNPs interact with serious illness, injury or assault to yourself	4

Figure S3: GWEIS Manhattan plot of SNPs interact with serious illness, injury or assault to a close relative	4

Figure S4: GWEIS Manhattan plot of SNPs interact with death of a spouse	5

Figure S5: GWEIS Manhattan plot of SNPs interact with marital separation/divorce	5

Figure S6: GWEIS Manhattan plot of SNPs interact with financial difficulties	6

Figure S7: GWEIS Manhattan plot of SNPs interact with addiction	6

Figure S8: GWEIS Manhattan plot of SNPs interact with time spent in computer game	7

GWEIS Plots for GAD-only

Figure S9: GWEIS Manhattan plot of SNPs interact with emotional abuse	7

Figure S10: GWEIS Manhattan plot of SNPs interact with sexual abuse	8

Figure S11: GWEIS Manhattan plot of SNPs interact with physical abuse	8

Figure S12: GWEIS Manhattan plot of SNPs interact with physical neglect	9

Figure S13: GWEIS Manhattan plot of SNPs interact with time spent in computer	9

Figure S14: GWEIS Manhattan plot of SNPs interact with serious illness, injury or assault to yourself	10

Figure S15: GWEIS Manhattan plot of SNPs interact with serious illness, injury or assault to a close relative	10

Figure S16: GWEIS Manhattan plot of SNPs interact with financial difficulties	11

GWEIS Plots for comorbid group

Figure S17: GWEIS Manhattan plot of SNPs interact with adoption	11

Figure S18: GWEIS Manhattan plot of SNPs interact with time spent in computer	12

Figure S19: GWEIS Manhattan plot of SNPs interact with death of spouse	12

Figure S20: GWEIS Manhattan plot of SNPs interact with maternal smoking around birth	13

Figure S21: GWEIS Manhattan plot of SNPs interact with addiction	13

Figure S21: GWEIS Manhattan plot of SNPs interact with marital separation/divorce	14

Figure S21: GWEIS Manhattan plot of SNPs interact with financial difficulties	14

Figure S1. Flowchart

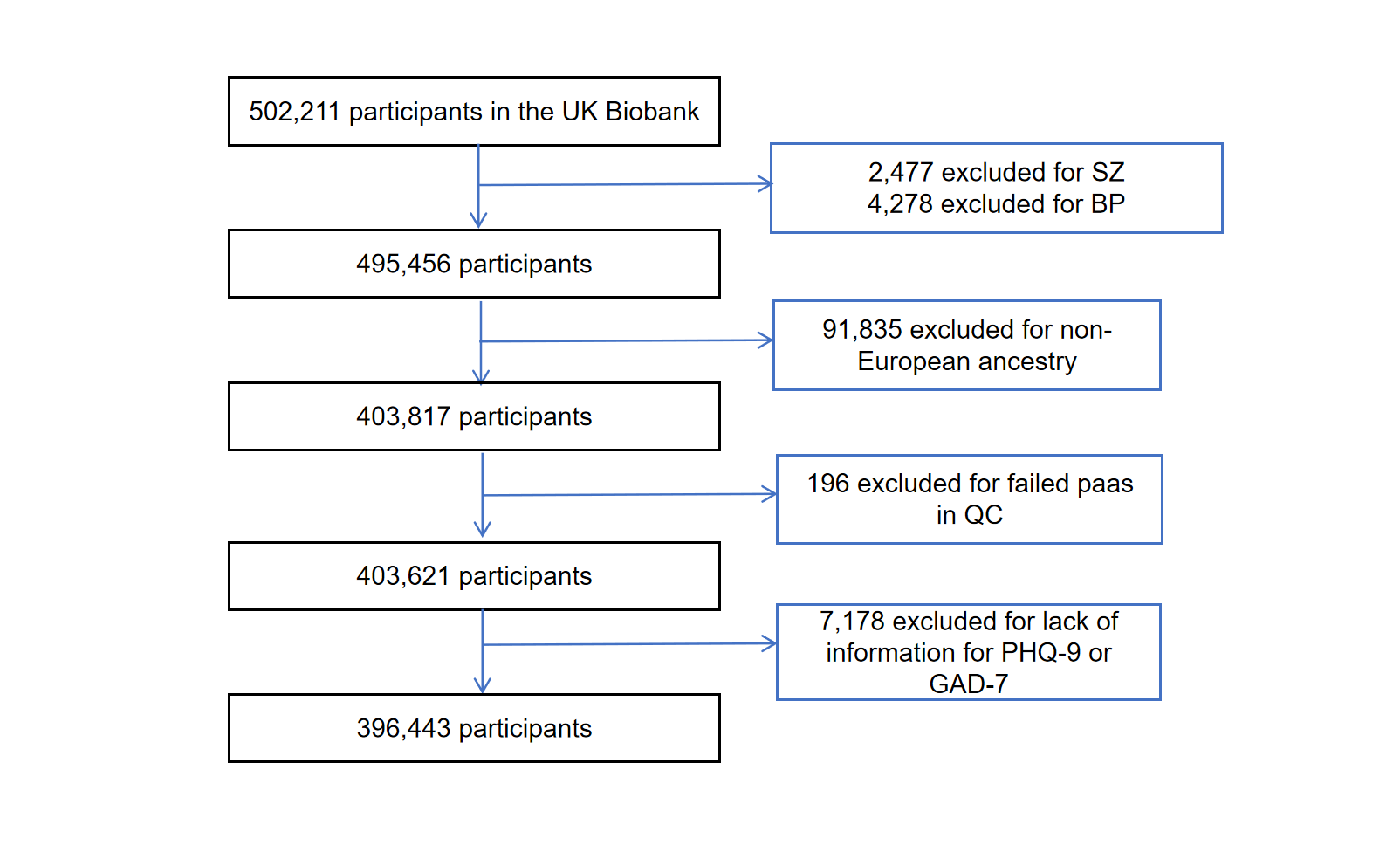

Note: BP: Bipolar disorder; GAD-7: the 7-item General Anxiety Disorder; PHQ-9: the 9-item Patient Health Questionnaire; SZ: Schizophrenia.

Figure S2. GWEIS Manhattan plot of SNPs interact with serious illness, injury or assault to yourself

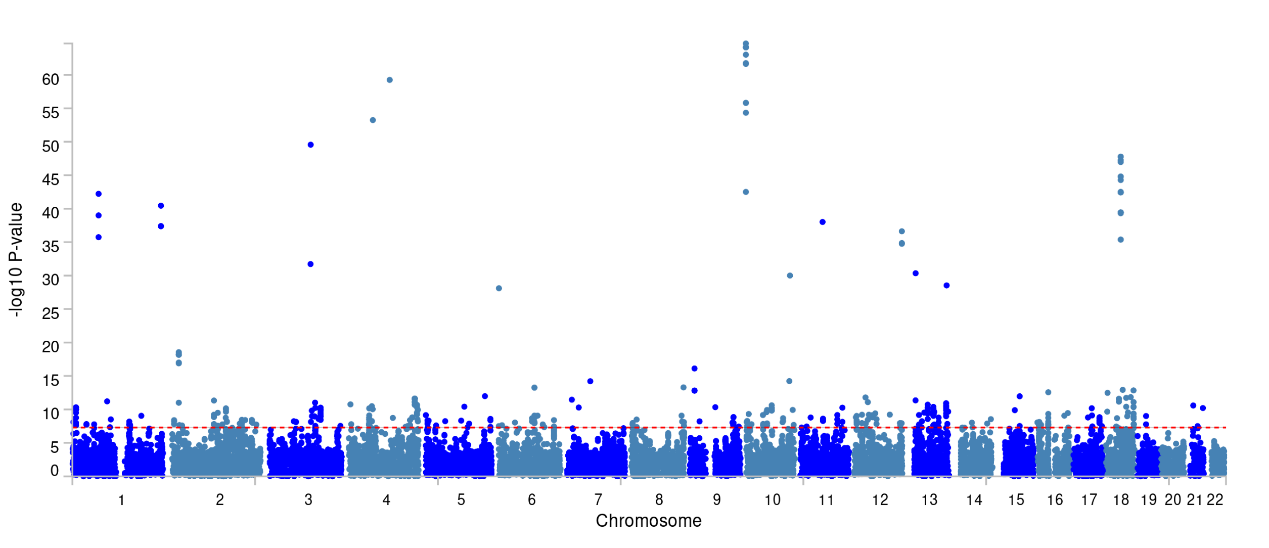

Note: Manhattan plot showing the -log10 p-value from GWEIS. The p-value is 5×10^-8^ indicating by red dotted line

Figure S3. GWEIS Manhattan plot of SNPs interact with serious illness, injury or assault of a close relative

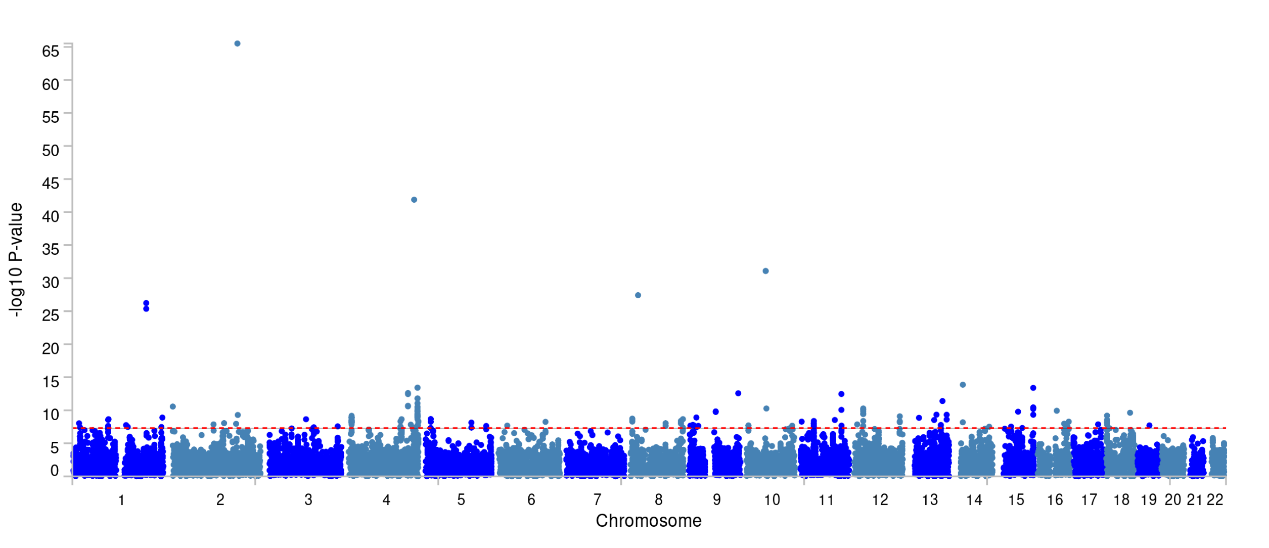

Note: Manhattan plot showing the -log10 p-value from GWEIS. The p-value is 5×10^-8^ indicating by red dotted line

Figure S4. GWEIS Manhattan plot of SNPs interact with death of a spouse

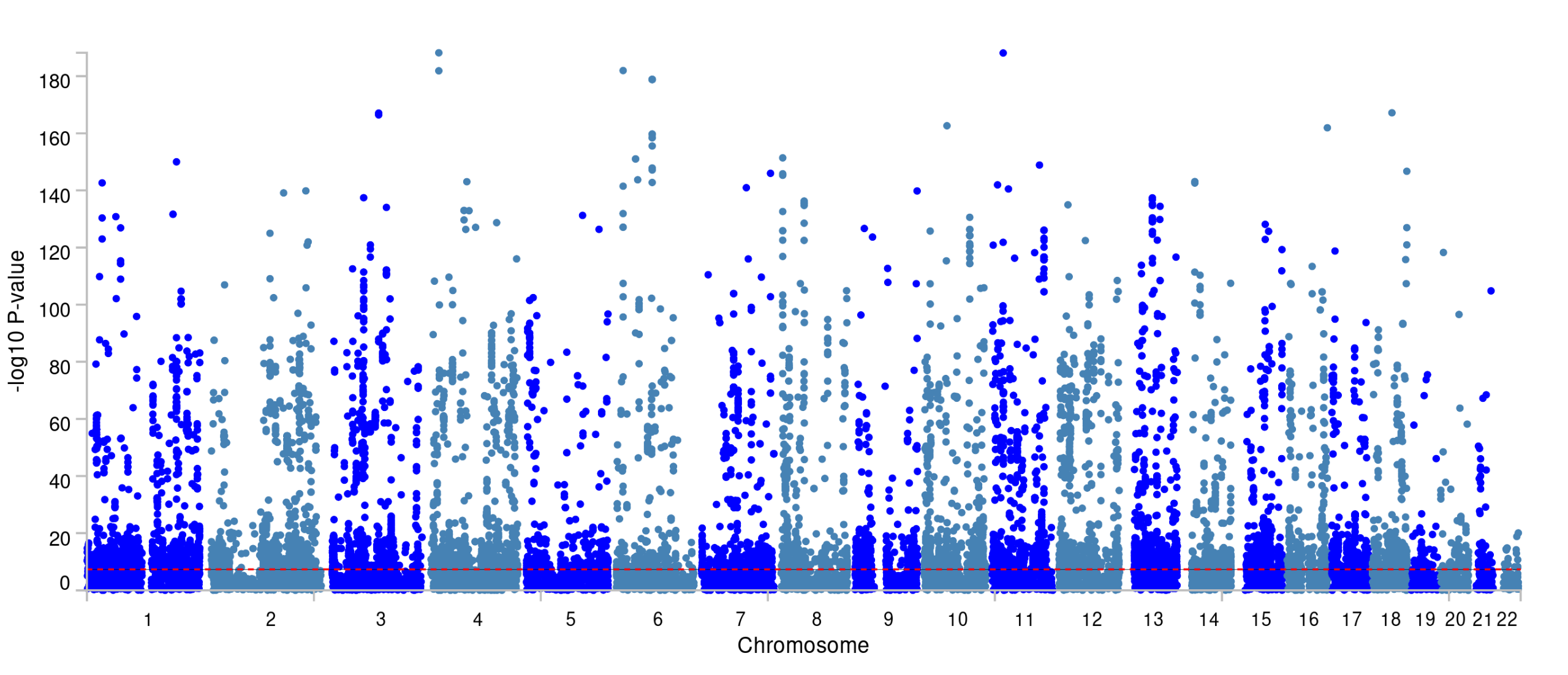

Note: Manhattan plot showing the -log10 p-value from GWEIS. The p-value is 5×10^-8^ indicating by red dotted line

Figure S5. GWEIS Manhattan plot of SNPs interact with marital separation/divorce

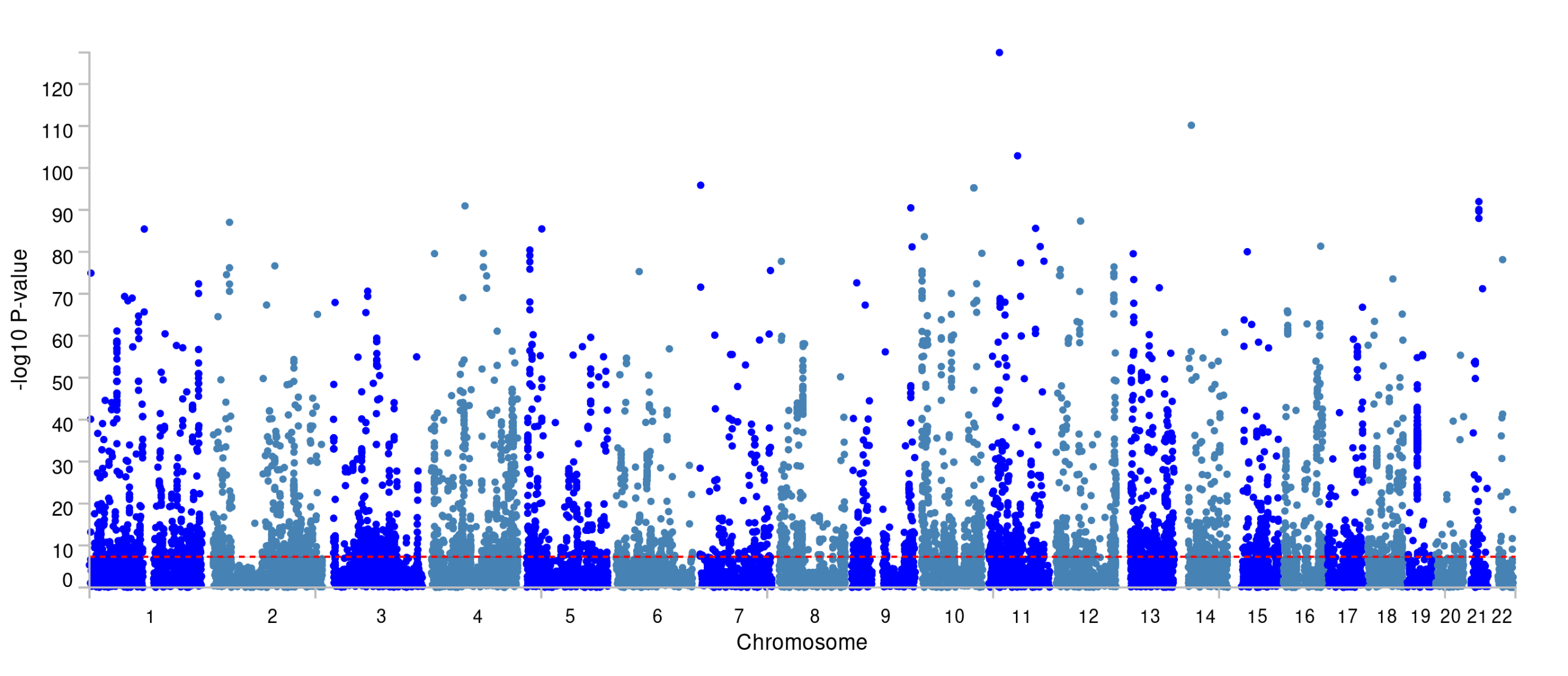

Note: Manhattan plot showing the -log10 p-value from GWEIS. The p-value is 5×10^-8^ indicating by red dotted line

Figure S6. GWEIS Manhattan plot of SNPs interact with financial difficulties

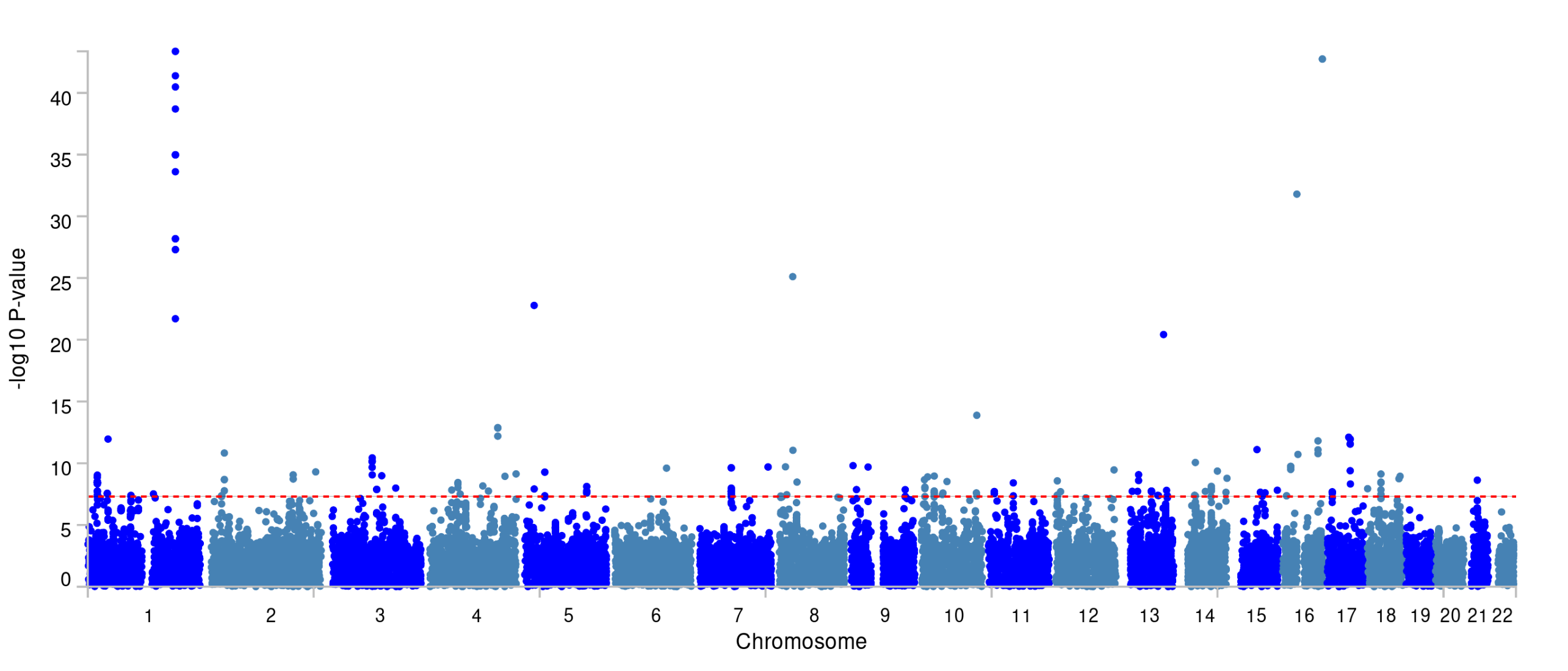

Note: Manhattan plot showing the -log10 p-value from GWEIS. The p-value is 5×10^-8^ indicating by red dotted line

Figure S7. GWEIS Manhattan plot of SNPs interact with addiction

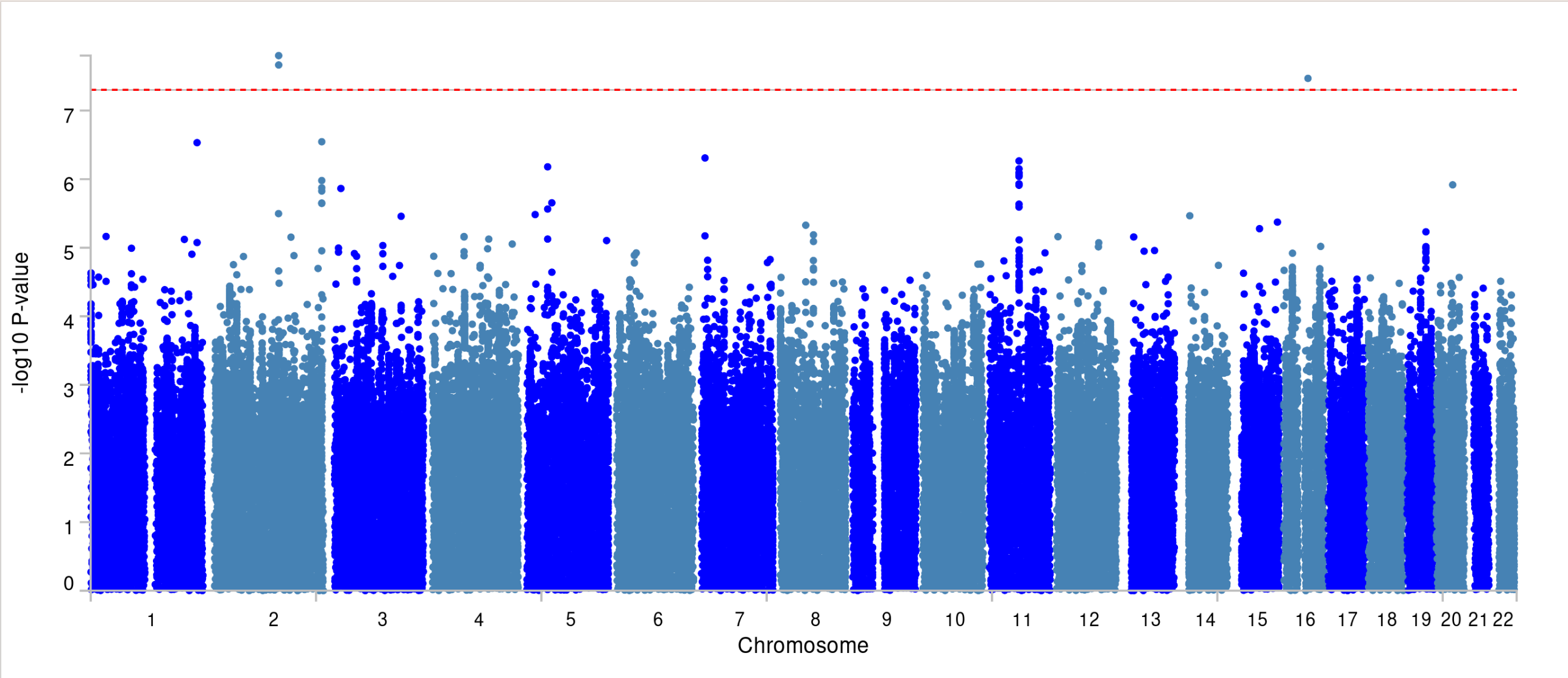
 Note: Manhattan plot showing the -log10 p-value from GWEIS. The p-value is 5×10^-8^ indicating by red dotted line

Figure S8. GWEIS Manhattan plot of SNPs interact with time spent in computer game

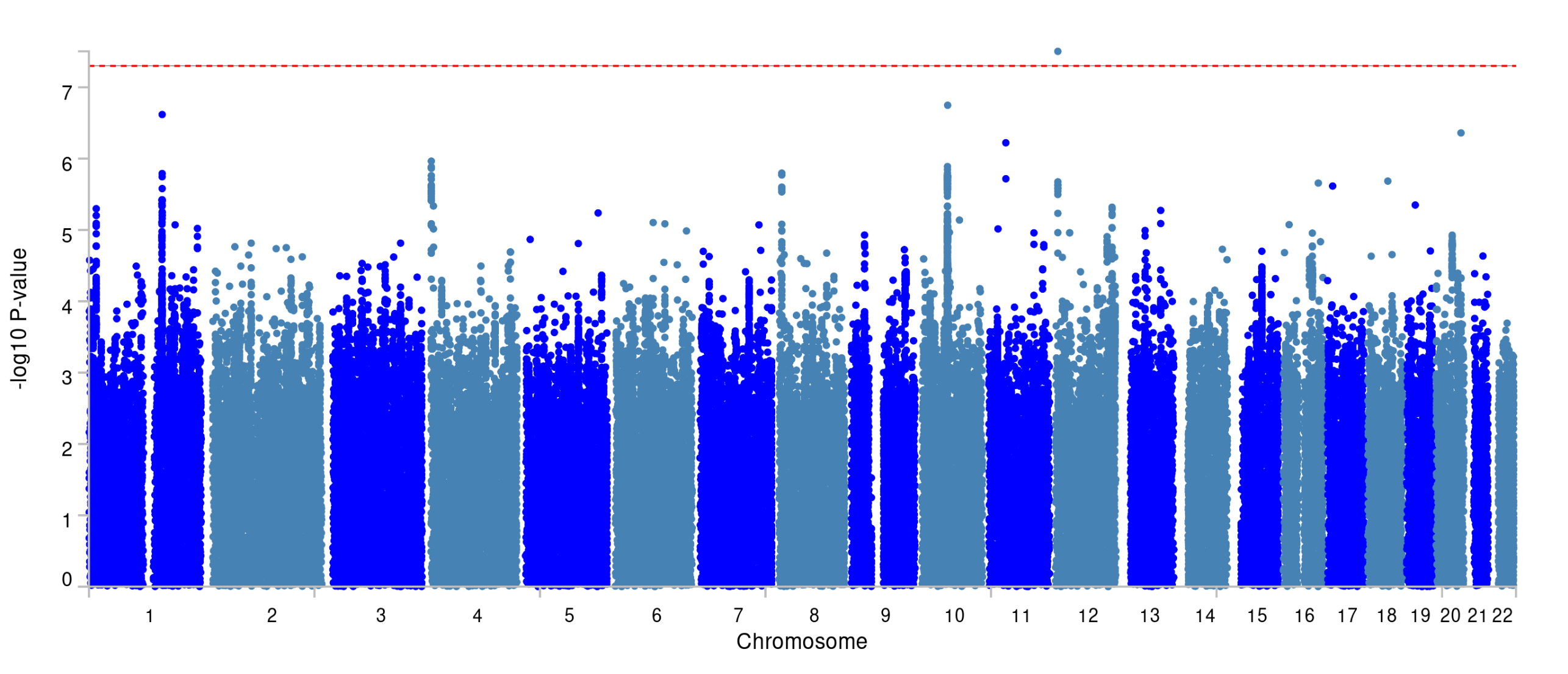

Note: Manhattan plot showing the -log10 p-value from GWEIS. The p-value is 5×10^-8^ indicating by red dotted line

Figure S9. GWEIS Manhattan plot of SNPs interact with emotional abuse

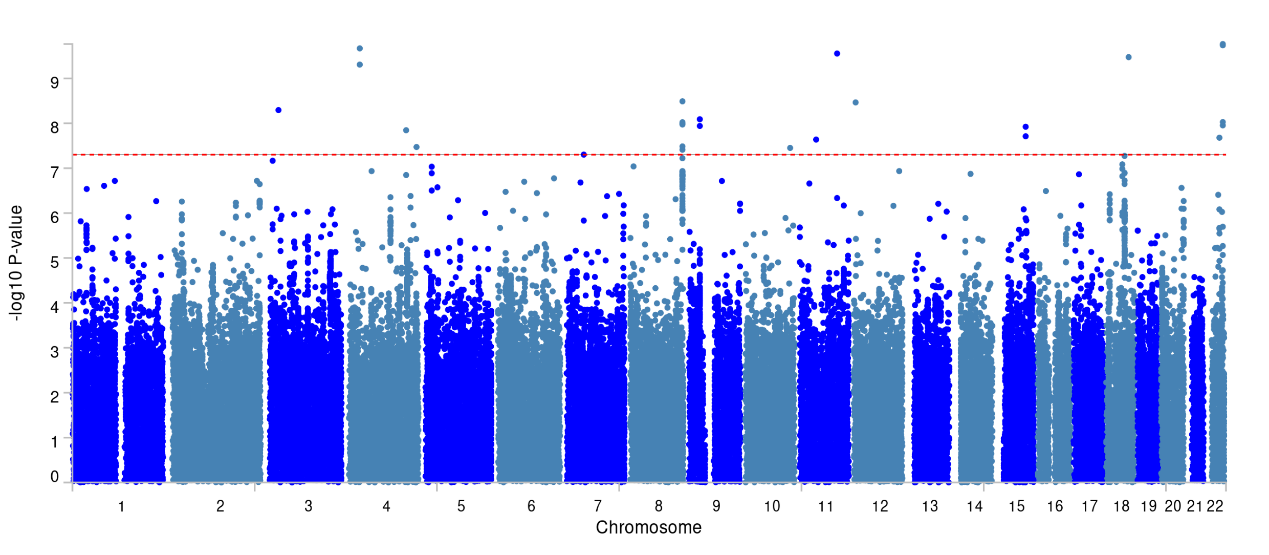

Note: Manhattan plot showing the -log10 p-value from GWEIS. The p-value is 5×10^-8^ indicating by red dotted line

Figure S10. GWEIS Manhattan plot of SNPs interact with sexual abuse

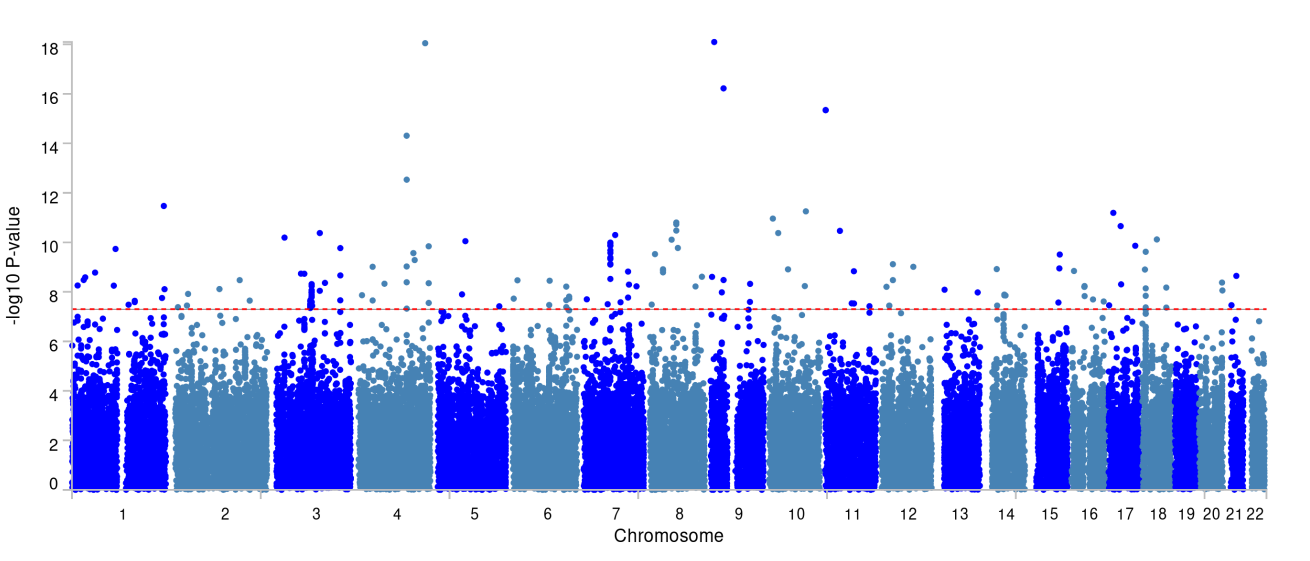

Note: Manhattan plot showing the -log10 p-value from GWEIS. The p-value is 5×10^-8^ indicating by red dotted line

Figure S11. GWEIS Manhattan plot of SNPs interact with physical abuse

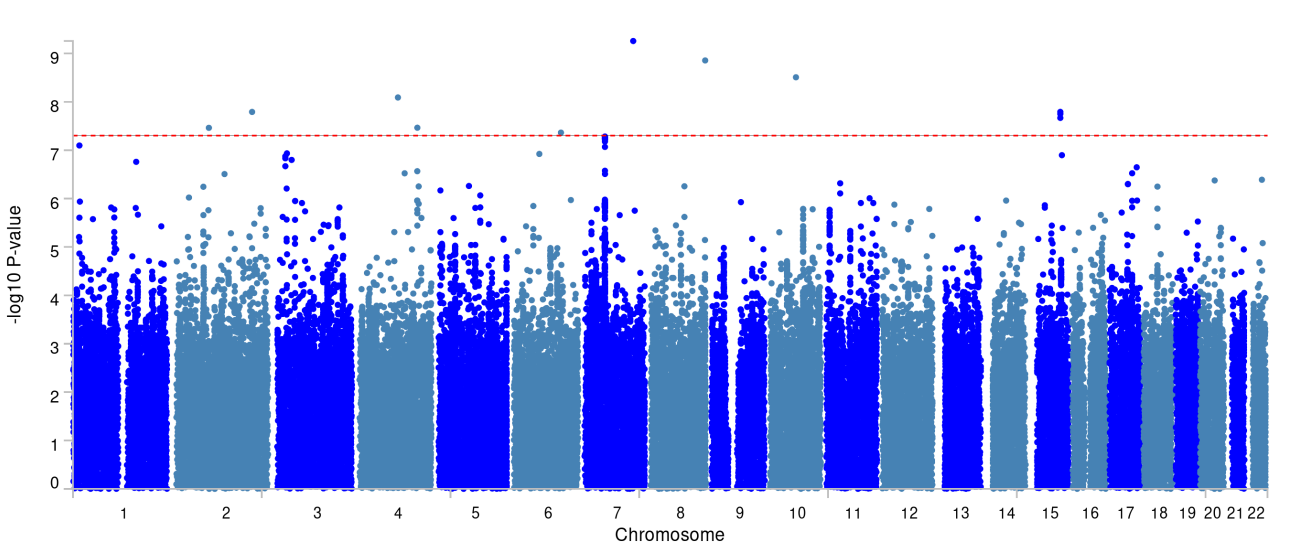

Note: Manhattan plot showing the -log10 p-value from GWEIS. The p-value is 5×10^-8^ indicating by red dotted line

Figure S12. GWEIS Manhattan plot of SNPs interact with physical neglect

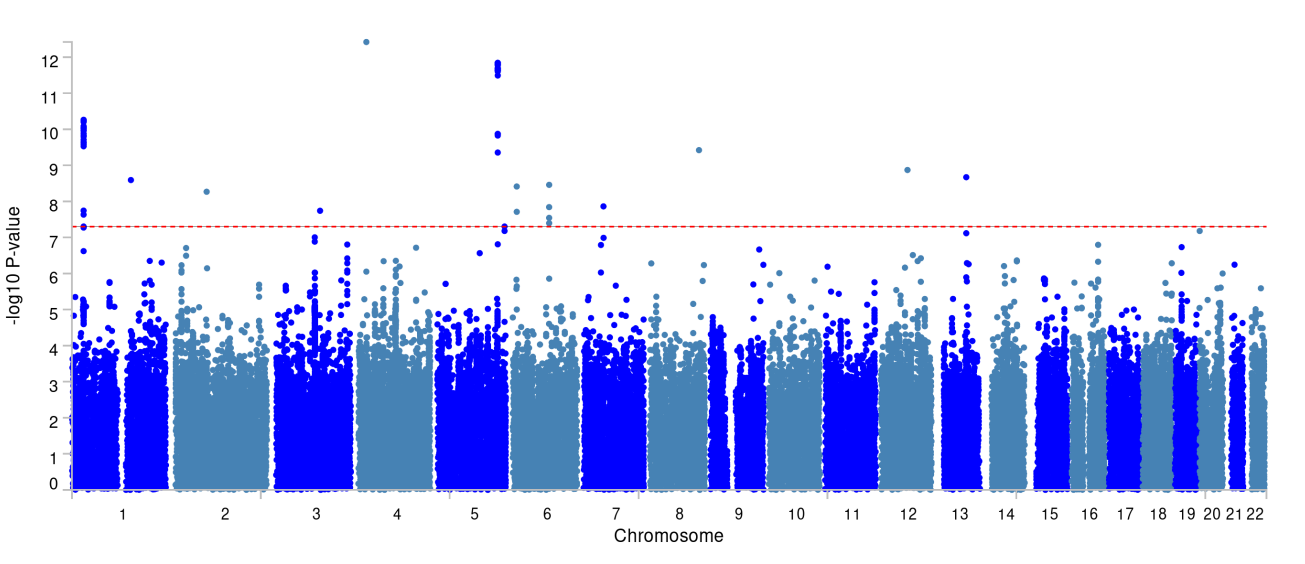

Note: Manhattan plot showing the -log10 p-value from GWEIS. The p-value is 5×10^-8^ indicating by red dotted line

Figure S13. GWEIS Manhattan plot of SNPs interact with time spent in computer

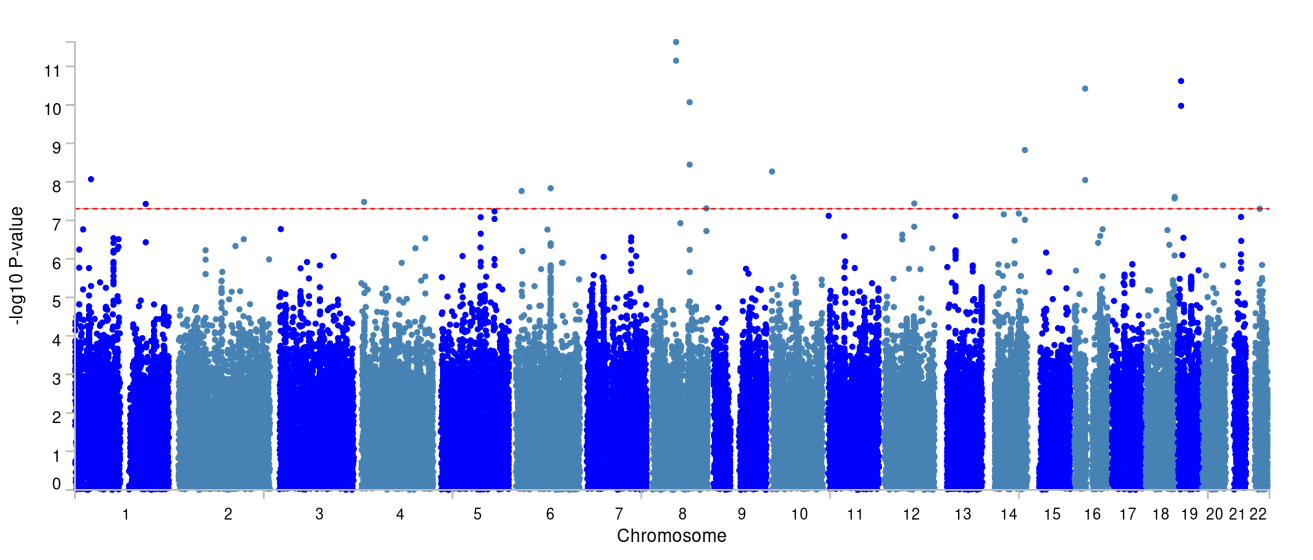

Note: Manhattan plot showing the -log10 p-value from GWEIS. The p-value is 5×10^-8^ indicating by red dotted line

Figure S14. GWEIS Manhattan plot of SNPs interact with serious illness, injury or assault to yourself

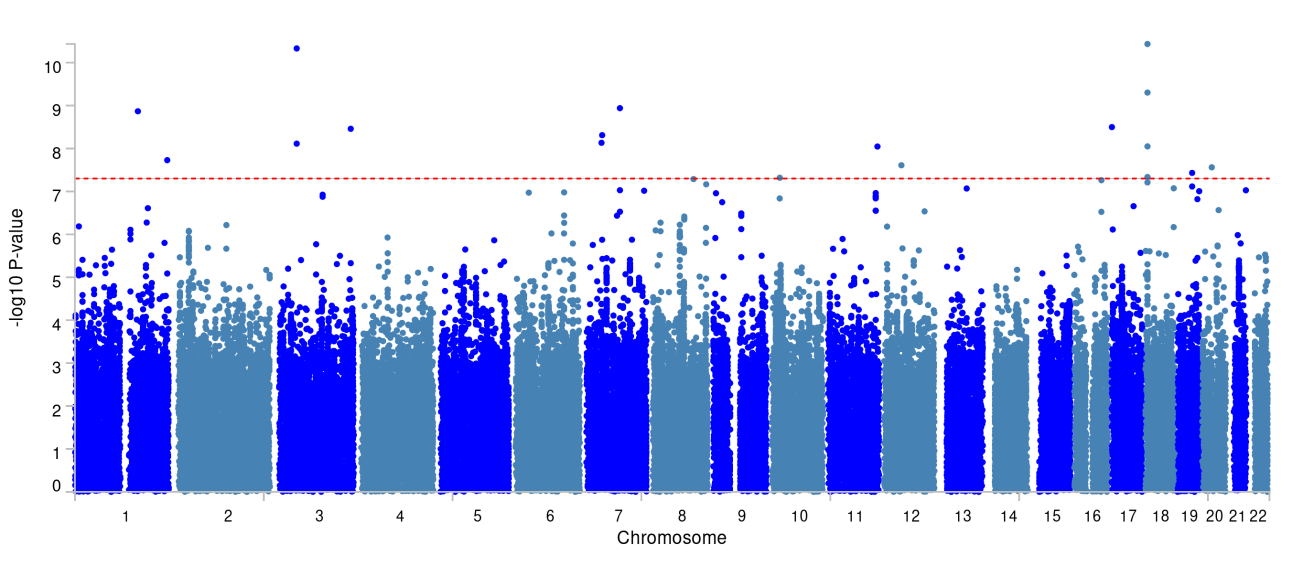

Note: Manhattan plot showing the -log10 p-value from GWEIS. The p-value is 5×10^-8^ indicating by red dotted line

Figure S15. GWEIS Manhattan plot of SNPs interact with serious illness, injury or assault to close relative

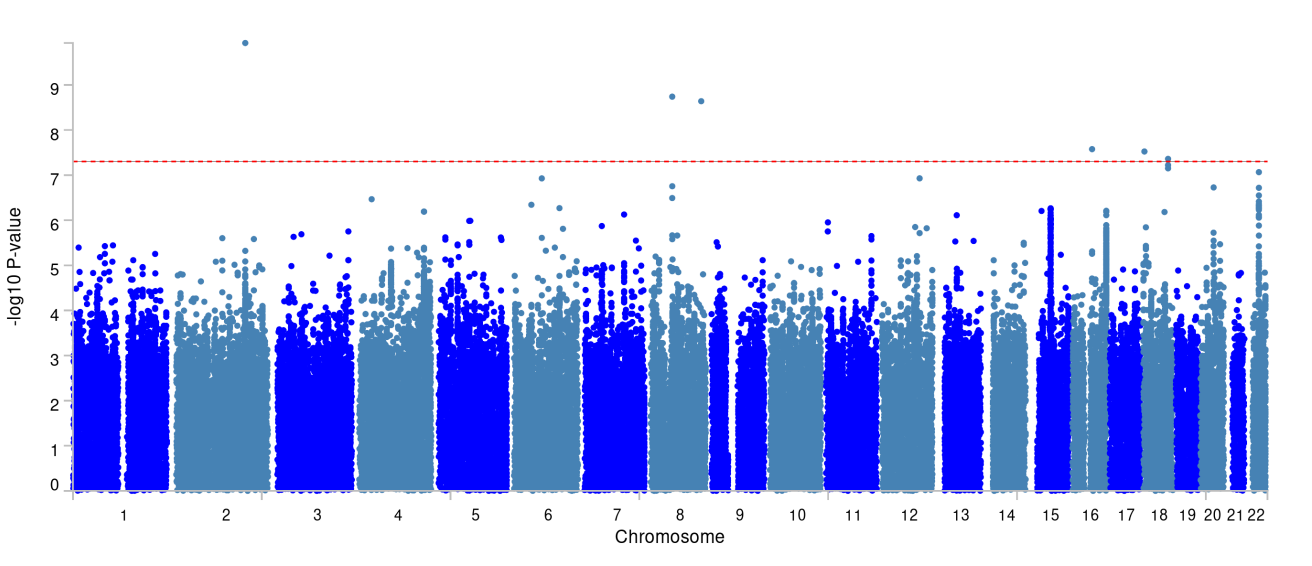

Note: Manhattan plot showing the -log10 p-value from GWEIS. The p-value is 5×10^-8^ indicating by red dotted line

Figure S16. GWEIS Manhattan plot of SNPs interact with financial difficulties

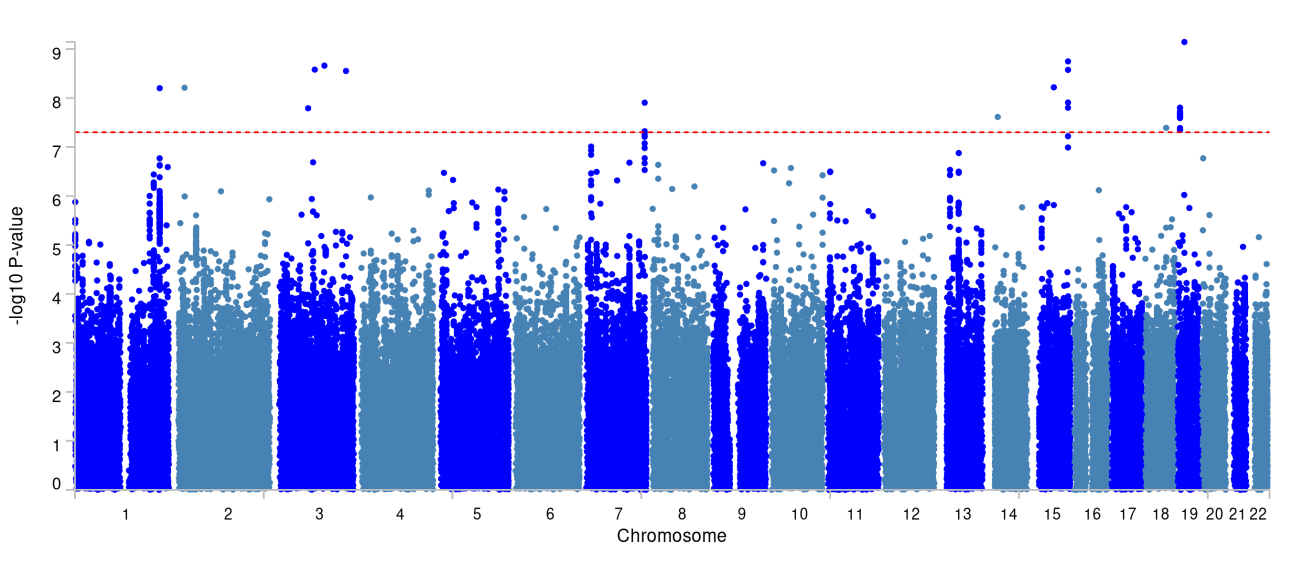

Note: Manhattan plot showing the -log10 p-value from GWEIS. The p-value is 5×10^-8^ indicating by red dotted line

Figure S17. GWEIS Manhattan plot of SNPs interact with adoption

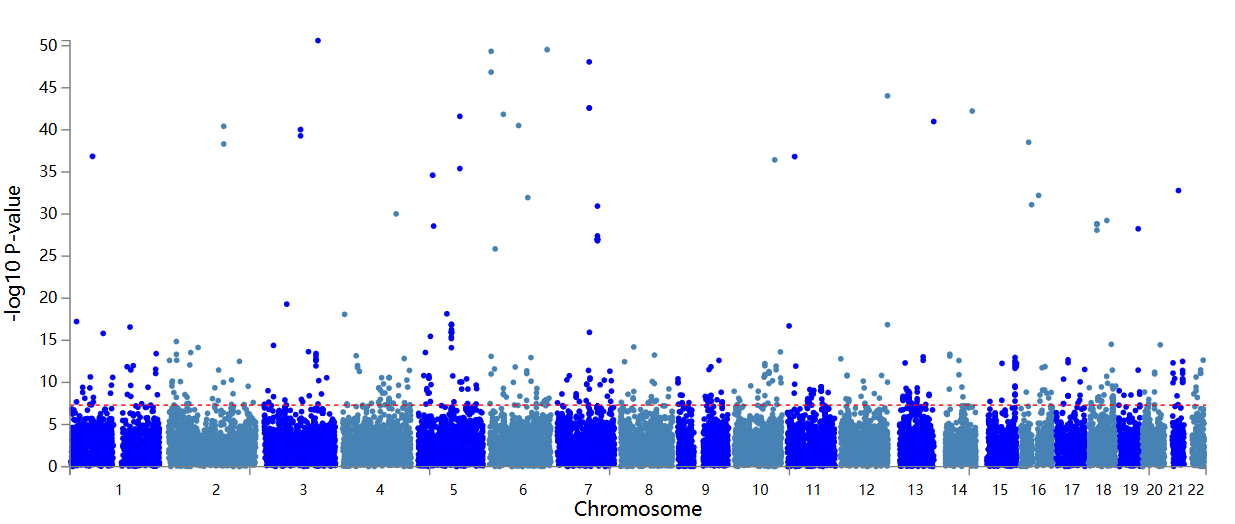

Note: Manhattan plot showing the -log10 p-value from GWEIS. The p-value is 5×10^-8^ indicating by red dotted line

Figure S18. GWEIS Manhattan plot of SNPs interact with time spent in computer

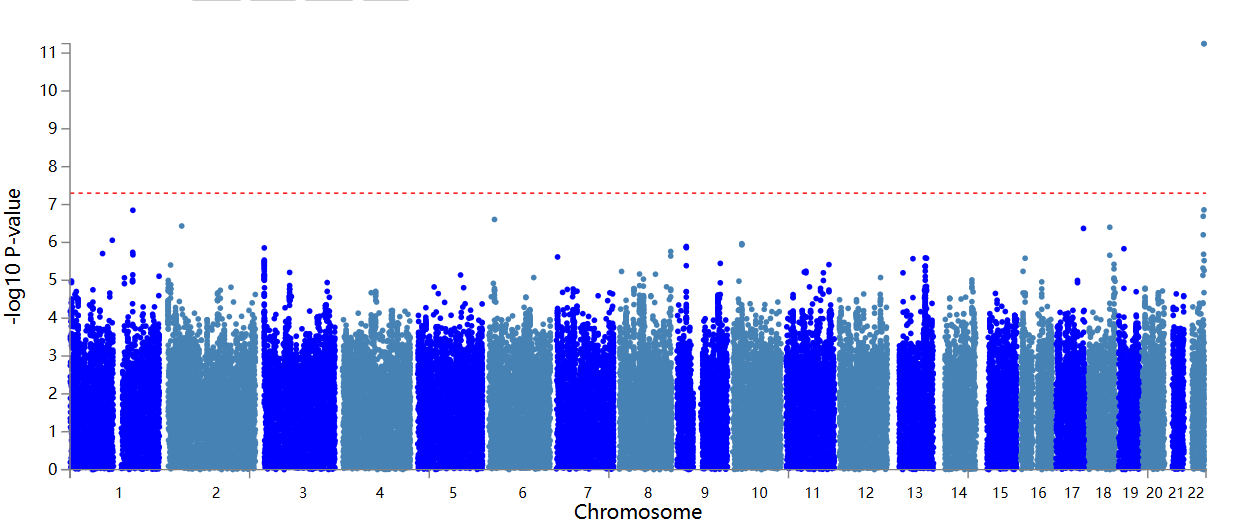

Note: Manhattan plot showing the -log10 p-value from GWEIS. The p-value is 5×10^-8^ indicating by red dotted line

Figure S19. GWEIS Manhattan plot of SNPs interact with death of spouse

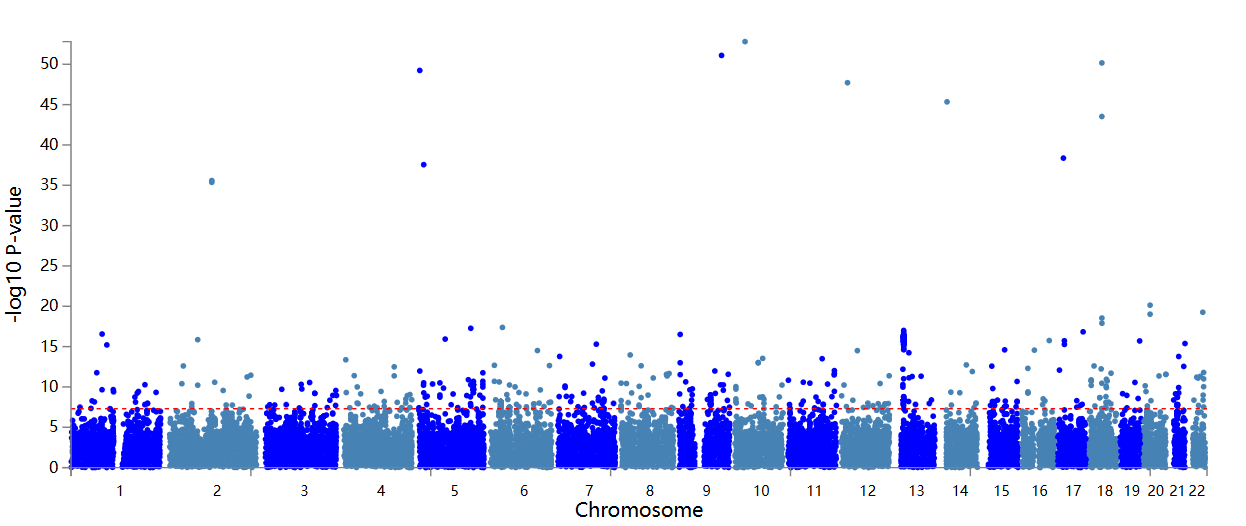

Note: Manhattan plot showing the -log10 p-value from GWEIS. The p-value is 5×10^-8^ indicating by red dotted line

Figure S20. GWEIS Manhattan plot of SNPs interact with maternal smoking around birth

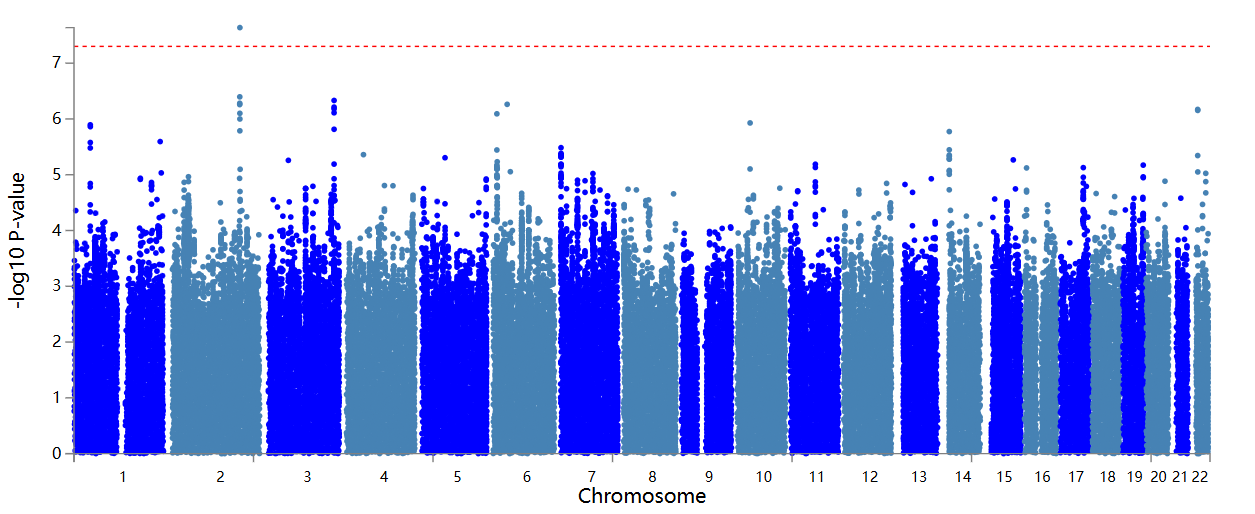

Note: Manhattan plot showing the -log10 p-value from GWEIS. The p-value is 5×10^-8^ indicating by red dotted line

Figure S21. GWEIS Manhattan plot of SNPs interact with addiction

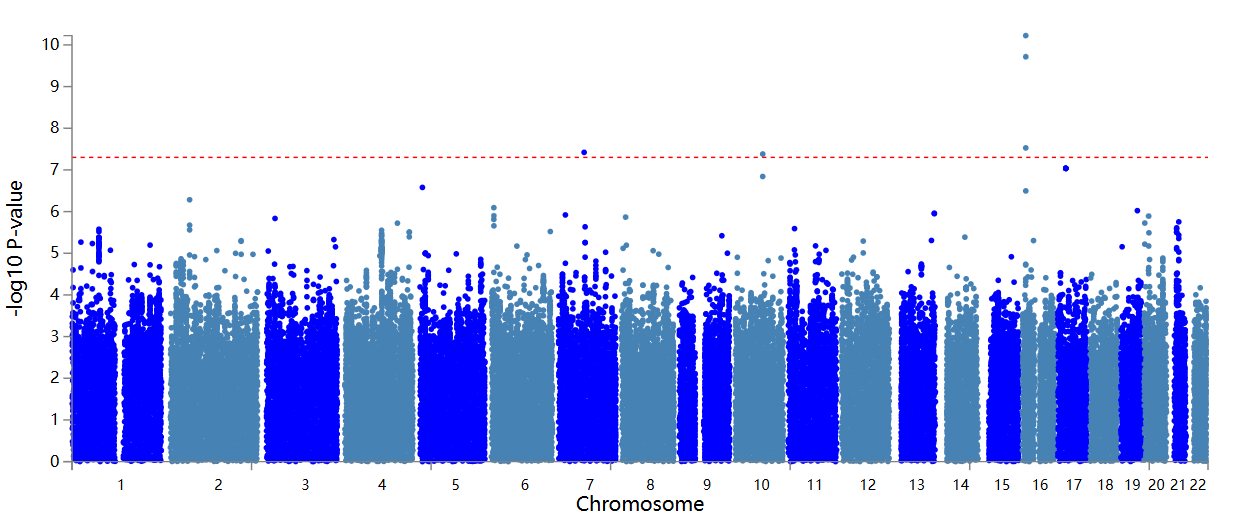

Note: Manhattan plot showing the -log10 p-value from GWEIS. The p-value is 5×10^-8^ indicating by red dotted line

Figure S22. GWEIS Manhattan plot of SNPs interact with marital separation/divorce

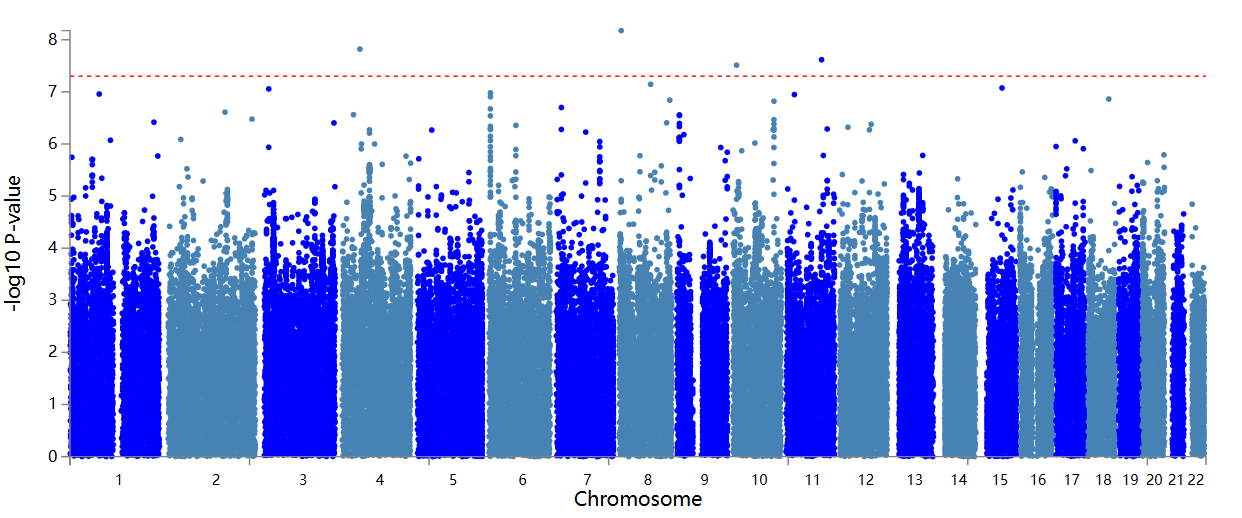

Note: Manhattan plot showing the -log10 p-value from GWEIS. The p-value is 5×10^-8^ indicating by red dotted line

Figure S23. GWEIS Manhattan plot of SNPs interact with financial difficulties

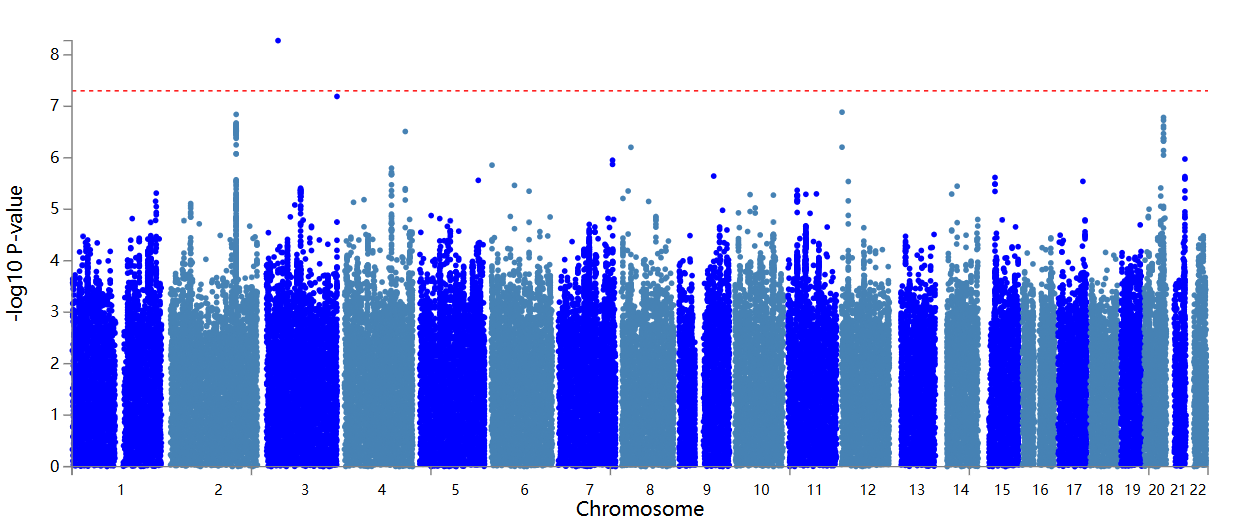

Note: Manhattan plot showing the -log10 p-value from GWEIS. The p-value is 5×10^-8^ indicating by red dotted line
